# Mapping generalizable brain-based depression subtypes across clinical, cognitive, and neurotransmitter dimensions

**DOI:** 10.64898/2026.06.25.26356577

**Authors:** Federica Colombo, Lidia Fortaner-Uyà, Tommaso Cazzella, Alfonso Martone, Camilla Monopoli, Cristina Colombo, Raffaella Zanardi, Matteo Carminati, Chiara Fabbri, Alessandro Serretti, Sara Poletti, Francesco Benedetti, Benedetta Vai

**Author notes:** Correspondence should be addressed to F.C.

## Abstract

Identifying generalizable brain-based biotypes across independent cohorts is critical for parsing heterogeneity in Major Depressive Disorder (MDD), yet robust subtypes spanning micro- and macroscales remain poorly defined. We applied stability-based clustering to cortical thickness data from 1,531 MDD individuals in UK Biobank (UKB), with external validation in 144 inpatients from IRCCS Ospedale San Raffaele (HSR). Two distinguishable clusters emerged (accuracy=87.5%), with one showing widespread cortical thinning, anergy-related symptoms, childhood trauma, and diabetes comorbidity. This profile generalized with 96.5% accuracy in a hold-out UKB sample and 80.6% in HSR. Mapping clusters’ cortical profiles onto Neurosynth meta-analytic activation patterns revealed a ventral-dorsal gradient linked with emotion regulation, interoceptive, and motivational processes. Spatial correlations with 19 neurotransmitter receptors and transporters obtained from positron emission tomography identified dopamine transporter as the dominant contributor in UKB, and histamine receptor H3 in HSR. These findings provide a reproducible framework linking MDD subtypes to multiscale biological complexity.

## Introduction

It is generally acknowledged that Major Depressive Disorder (MDD) is highly heterogeneous both in its etiology and clinical profiles^1,2^. With up to 30–40% of patients failing to respond to standard treatments^3^ and substantial excess mortality linked to suicide and physical comorbidities^4^, this heterogeneity challenges the effectiveness of one-size-fits-all treatment approaches, as current diagnostic frameworks fail to capture the varied underlying biological mechanisms^5^. In the context of precision medicine, the primary objectives are to determine how this multiscale complexity unfolds in patients with differing clinical outcomes and to identify common downstream mechanisms for these individuals^6,7^.

Making progress on these fronts inevitably brings the brain into focus, as it represents the final common pathway through which genetic, environmental, and psychosocial factors converge to produce the heterogeneous clinical manifestations of depression^8,9^. Among neuroimaging phenotypes, cortical thickness (CT) exhibits unique heritability profiles, close proximity to underlying cytoarchitectural properties^10^, and high test-retest reliability across scanner platforms^11^. Widespread cortical thinning in the frontal, temporal, cingulate, and insular regions has been consistently replicated in MDD^12,13^, mapping onto distinct symptom dimensions, including depression severity, melancholic features^14,15^, and impaired executive functions in treatment-resistant patients^16^.

Understanding the neurobiological heterogeneity of depression requires, however, moving beyond one single modality and integrate different spatial scales to comprehend how macroscale changes may be driven by microscale processes, which can identify potential innovative therapeutic targets^17^. Neurotransmitter systems represent one such bridge; acting on receptors bound to neurons’ membrane to drive intercellular signalling, they modulate neural dynamics and shape large-scale network communication, influencing behaviours^18,19^. They also affect the cortical structure, with cortical thinning in MDD spatially correlating with the density of dopaminergic, serotoninergic, and glutamatergic receptor densities^20,21^. Their translational relevance is further underscored by their role as primary targets of first-line antidepressant treatments, from selective serotonin reuptake inhibitors (SSRIs) and serotonin-norepinephrine reuptake inhibitors (SNRIs) acting on monoaminergic transporters^22^, to dopaminergic agents such as bupropion showing efficacy in patients unresponsive to first-line treatments^23,24^, as well as to treatments for treatment-resistant depression, such as ketamine, targeting N-methyl-D-aspartate (NMDA) receptors^25^, and emerging serotoninergic compounds such as psilocybin^26^.

Yet, the high rates of treatment resistance suggest that these systems are not uniformly disrupted across patients, and that individual differences in receptor architecture may uniquely shape both cortical alterations and cognitive/affective processes^27^. Shedding light on this puzzle, research in MDD is increasingly moving towards dimensional brain-based models capable of capturing subtype-specific signals in a data-driven way^16,28^. Studies focusing on resting-state measures have yielded anhedonia and anxiety-related subtypes with opposing patterns of functional connectivity^29,30^ and differential responses to antidepressant treatments^31^. Structural imaging has identified subtypes differing in white matter integrity, cognitive profiles^32,33^, and transcriptomic signatures^34^. Along with these findings, our previous work demonstrated that structural neuroimaging measures can differentiate between neurally preserved and compromised subtypes of MDD, with the latter being associated with an inflammatory burden (e.g., interleukin-6), symptoms of anergy, poor response to antidepressants, and childhood trauma ^35^.

Achieving clinically relevant subtying, however, requires subtypes that are robustly generalizable to different populations and clinical contexts. The reliance on small and unrepresentative samples is a persistent bottleneck in neuroimaging research, producing models with inflated predictive performance that frequently fail to generalize beyond the original study context ^36,37^. Beyond replication, meaningful subtyping requires data that are not only “broad” but also “deep” (spanning multiple levels of analysis), as the combination of both enables clinically useful models explaining the disorder’s multi-level complexity^38,39^. Yet even when such data are available, robust external validation in fully independent datasets remains rarely adopted, while validation against a hold-out subsample is still a common practice^40^.

Here, we addressed these gaps by leveraging a large community sample (N=1,531 from the UK Biobank, UKB) to uncover neurobiologically grounded subtypes of depression based on CT. By implementing a robust stability-based clustering approach, we rigorously validated the identified subtypes in both a hold-out UKB subsample and a fully independent external cohort of MDD inpatients. We then examined the cognitive and affective processes linked to clusters’ cortical signature and the neurochemical mechanisms driving them, using open-source positron emission tomography (PET) data from 1,200 healthy individuals. Finally, the clinical relevance of the clusters in terms of their association with distinct symptom profiles was reported (including depression severity, cardiometabolic comorbidities, and treatment resistance).

## Results

All analyses were embedded in a three-stage framework: i) model development in the 55% of UKB cohort (UKB training set, N=842); ii) internal validation in the remaining 45% (UKB hold-out test set, N=689); iii) external validation in the San Raffaele Hospital (HSR) clinical cohort (N=144). The 55-45 training-test splitting in UKB was stratified to ensure a balanced distribution of age and sex across splits, while full propensity score matching was applied in HSR to control for demographic differences between cohorts. Clusters identified in the UKB were also compared with healthy controls (HC, N=827). We then profiled clusters on clinically relevant outcomes and mapped the spatial distribution of their cortical differences onto meta-analytical functional activation maps (*Neurosynth*). Finally, we assessed the spatial covariance of these cortical signatures with neurotransmitter system distributions (*neuromaps*) to characterize their neurochemical correlates (Fig.1).

**Figure 1.**
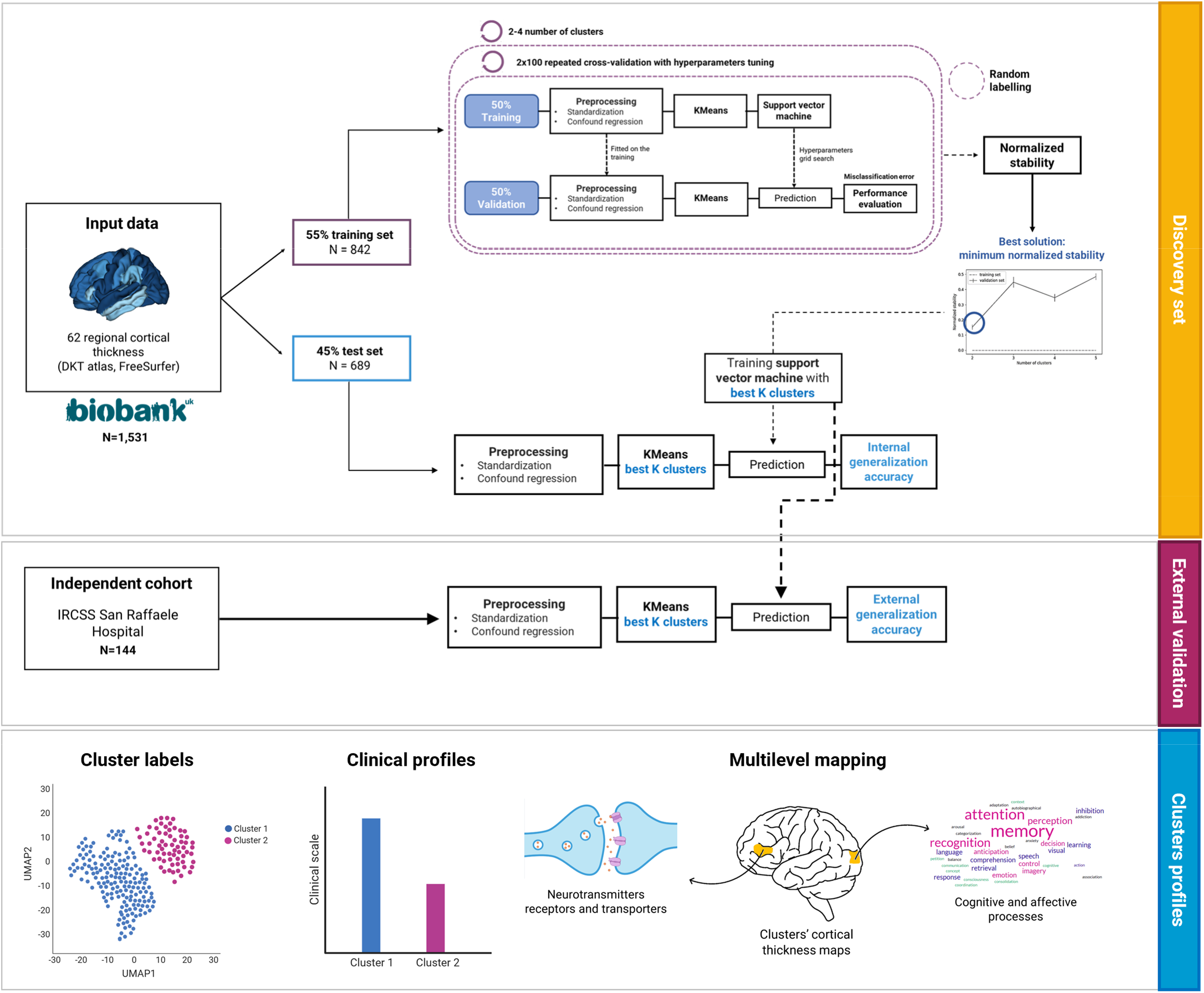
Graphical representation of the analytical pipeline. Region-wise mean CT values (DKT atlas) were used as input features for stability-based clustering (*NeuReval*), combining KMeans clustering with SVM classification. The optimal clustering solution was selected by minimizing normalized stability across 100X2-fold repeated cross-validation in the UKB training set (55%). The optimal solution was then applied to the UKB held-out set (45%) and to the independent HSR inpatient cohort, where the SVM model trained on UKB was used to predict cluster labels. Model significance was assessed via 10,000 permutations. The identified clusters were then profiled for outcome variables. Between-cluster CT differences were quantified as regional Cohen’s d maps for each cohort, serving as input for neurotransmitter receptor/transporter and cognitive function mapping.

### Sample characteristics

In UKB, patients’ characteristics were similar between sets. Compared to HC, MDD patients showed higher body mass index (BMI), frequency of any diabetes comorbidity (Data-Field 2443), and exposure to childhood trauma. Comparing UKB and HSR cohorts for common variables, patients in UKB were older and with higher BMI, age of onset, and frequency of depression with atypical features compared to HSR patients. Conversely, higher percentages of treatment-resistant patients were reported in HSR cohort compared to UKB (Supplementary Table S1-S3).

### Identification of brain-based subtypes in UKB

Using the UKB training set (N=842), two clusters were identified as the best clustering solution, with a cross-validation accuracy of 87.5% (p<0.001) (Supplementary Table S4). Cluster 1 (N=386) was characterized by widespread cortical thinning relative to Cluster 2 (N=456), with the highest effect sizes observed in the bilateral frontal and parietal regions, alongside additional thinning in the temporal, insular, and sensorimotor cortices (*d* = 0.48-1.65) (Fig.2a). Compared to HC, Cluster 1 showed lower global CT than both Cluster 2 and HC, whereas HC displayed an intermediate profile between the two clusters (Supplementary Fig.S1, Table S5).

**Figure 2.**
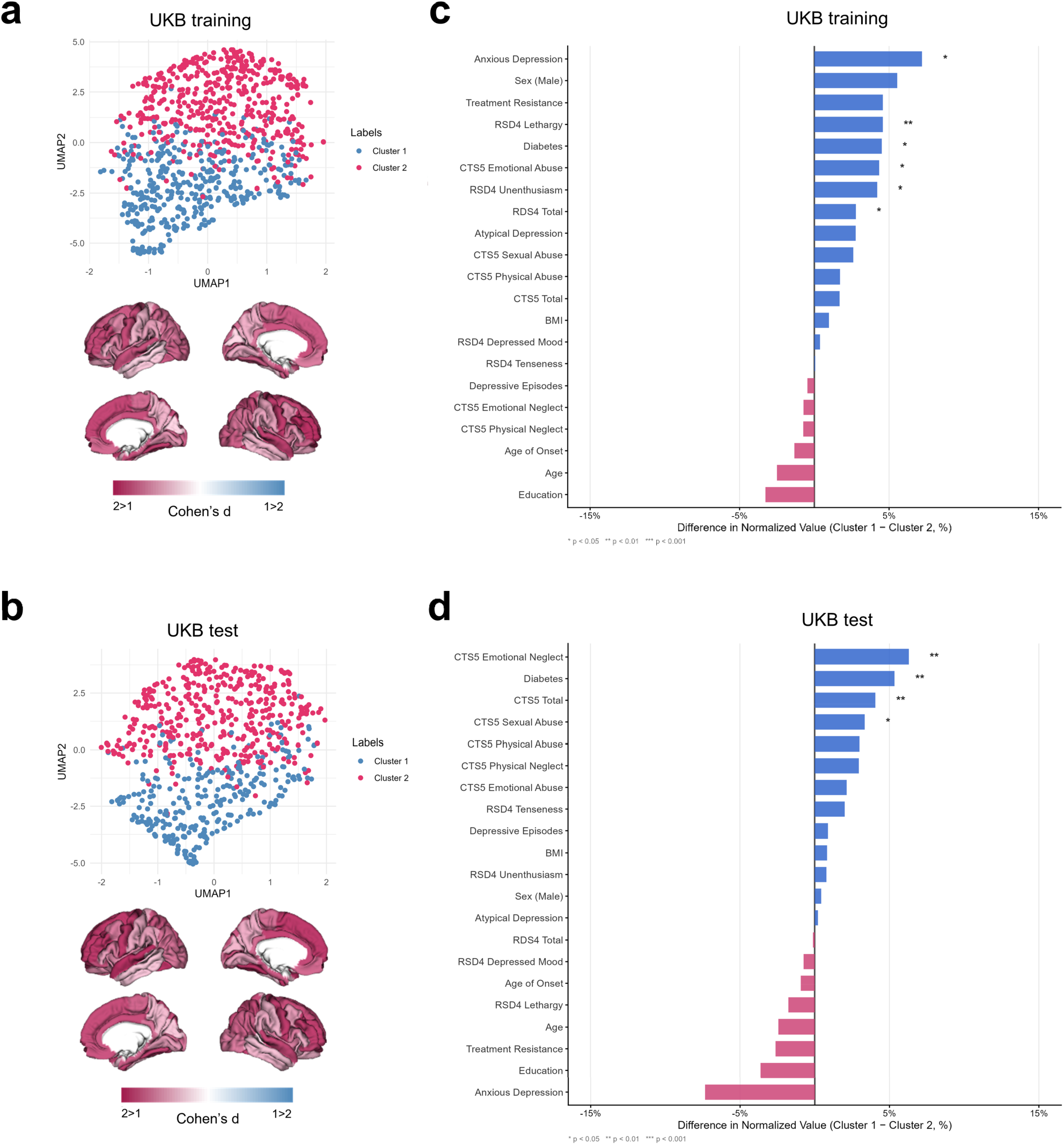
Results of stability-based relative clustering validation in the UKB. a-b, Plot of UMAP-reduced data input into the clustering for the UKB training set (a) and UKB test (b). The individuals are colored by subtype over the density plots. Cohen’s d brain maps describing the magnitude of differences in cortical thickness between clusters are represented on cortical surfaces. Positive values (pink) indicate higher values for Cluster 2 compared to Cluster 1, whereas negative values (blue) indicate higher values for Cluster 1 compared to Cluster 2. c-d, Differences in clinical and demographic characteristics between MDD clusters identified in the UKB training set (c) and test set (d). Bars represent the difference in normalized values (0–100 scale) between Cluster 1 and Cluster 2 for each variable. Blue bars indicate variables with higher normalized mean values in Cluster 1; pink bars indicate variables with higher normalized mean values in Cluster 2. Continuous variables were normalized to a 0–100 scale based on the global minimum and maximum across both clusters; binary variables are expressed as the percentage of individuals with the characteristic. Significance levels are based on p-values. UKB. UK Biobank; UMAP, Uniform Manifold Approximation and Projection for Dimension Reduction; BMI, body mass index; RDS4, Rating of Depression Scale-4; CTS5, Childhood Trauma Screener-5.

This solution generalized with 96.5% accuracy in the hold-out UKB test set (N=689) (p<0.001). Cluster 1 (N=279) again showed less preserved brain structure than Cluster 2 (N=410), with the largest effect sizes located in the bilateral fronto-parietal regions (*d*=0.38-1.60) (Fig.2b). Comparing clusters with HC, the same pattern observed in the training set was replicated, with HC falling between the two clusters in terms of global and regional CT (Supplementary Fig.S1, Table S6).

We then explored associations with outcome variables. In the UKB training set, both clusters showed higher Recent Depressive Symptoms scale (RDS-4) and Childhood Trauma Screener (CTS) total scores compared to HC, alongside higher BMI (q<0.001). Cluster 1 was distinguished from Cluster 2 by greater RDS-4 lethargy and unenthusiasm (q=0.005 and q=0.023), CTS emotional abuse (q=0.018), and prevalence of diabetes (q=0.018). (Fig.2c, Supplementary Table S7).

A broadly consistent profile was observed in the UKB hold-out test set. Both clusters showed higher RDS-4 total scores, childhood trauma, and BMI compared to HC (q<0.001). Cluster 1 was additionally characterized, relative to Cluster 2, by higher overall childhood trauma (q=0.007), including emotional neglect (q=0.008) and sexual abuse (q=0.025), and a higher prevalence of diabetes compared to both HC and Cluster 2 (q<0.001) (Fig.2d, Supplementary Table S8).

### External generalizability of the brain-based subtypes in HSR cohort

The 2-clusters solution replicated in an independent clinical cohort of MDD inpatients (HSR) with 80.6% accuracy (p=0.002). Most patients (80.6%, N=116) were assigned to Cluster 1, which showed more pronounced cortical thinning than Cluster 2 (N=28), consistent with the UKB findings. This pattern was particularly evident in the bilateral fronto-temporo-parietal regions and extended to the cingulate and posterior cortices (*d*=0.35-3.21) (Fig.3a, Supplementary Table S9). When comparing clusters for outcome variables, no statistical analyses survived multiple comparisons. A trend toward significance emerged for the association of Cluster 1 with treatment resistance (p=0.07) and comorbidity with cardiometabolic disorders (p=0.07). This cluster was also associated with older age (p=0.02) and age of onset (p=0.02) (Fig.3b, Supplementary Table S10).

**Figure 3.**
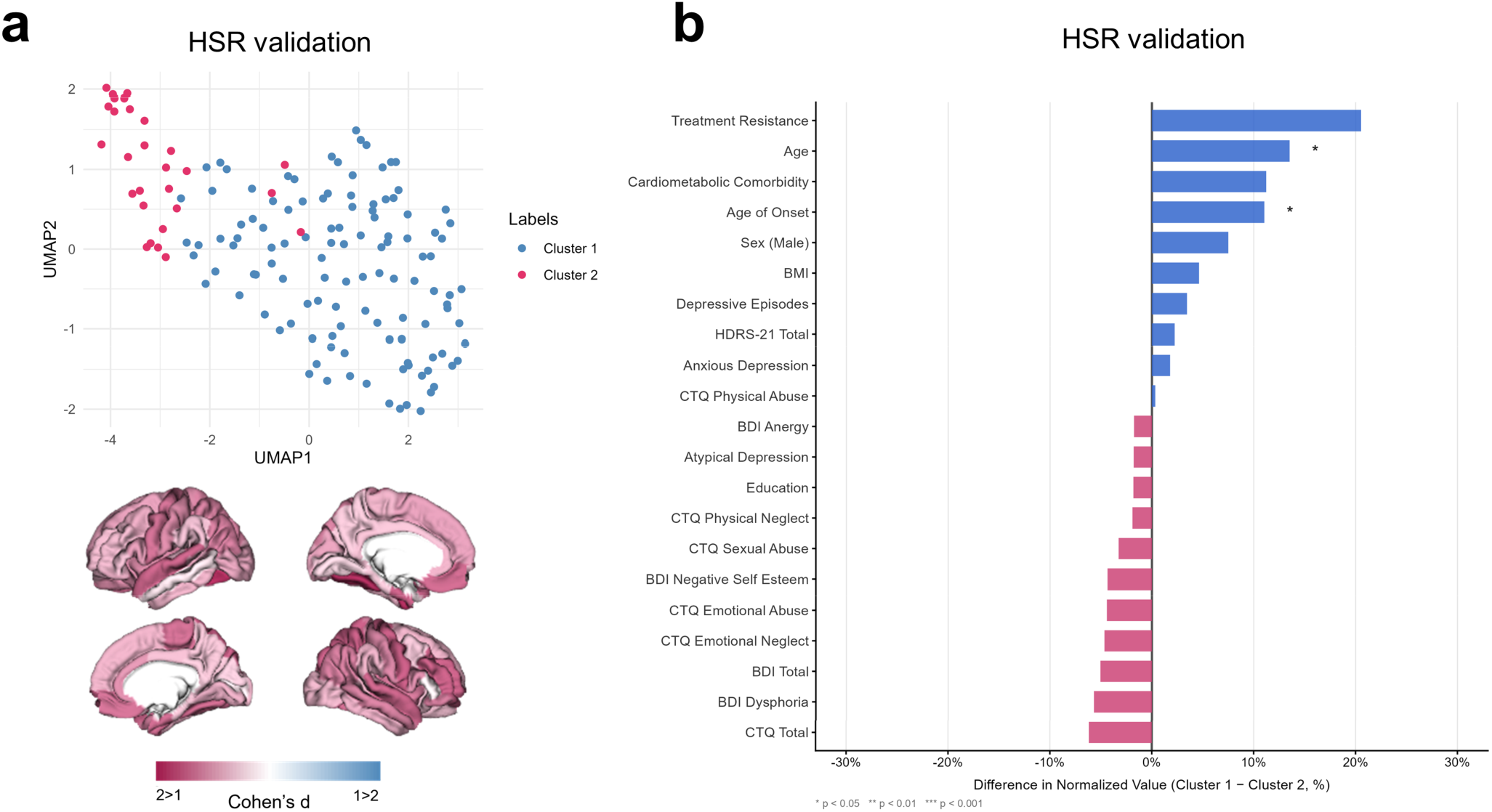
External validation of UKB stratification model in HSR. a, Plot of UMAP-reduced data input into the clustering for the HSR validation set. The individuals are colored by subtype over the density plots. Cohen’s d brain maps describing the magnitude of differences in cortical thickness between clusters are represented on cortical surfaces. Positive values (pink) indicate higher values for Cluster 2 compared to Cluster 1, whereas negative values (blue) indicate higher values for Cluster 1 compared to Cluster 2. b, Differences in clinical and demographic characteristics between MDD clusters identified in the HSR cohort. Bars represent the difference in normalized values (0–100 scale) between Cluster 1 and Cluster 2 for each variable. Blue bars indicate variables with higher normalized mean values in Cluster 1; pink bars indicate variables with higher normalized mean values in Cluster 2. Continuous variables were normalized to a 0–100 scale based on the global minimum and maximum across both clusters; binary variables are expressed as the percentage of individuals with the characteristic. Significance levels are based on raw p-values. HSR, San Raffaele Hospital; UMAP, Uniform Manifold Approximation and Projection for Dimension Reduction; BMI, body mass index; HDRS-21, Hamilton Depression Rating Scale - 21 items; BDI, Beck Depression Inventory; CTQ, Childhood Trauma Questionnaire.

### Mapping cognitive function to clusters’ cortical profiles

Next, we examined whether the spatial distribution of clusters’ cortical differences corresponded to the brain signatures of cognitive/affective processes derived from *Neurosynth* functional activation maps. In the UKB training set, Partial Least Squares (PLS) analysis extracted a significant latent variable (LV) associating clusters’ cortical profile to whole-brain functional activations (p_spin_=0.036, r=0.62). This association remained significant even with distance-dependent cross-validation, confirming the robustness of the association (mean out-of-sample r(60)=0.40, p_spin_=0.003) (Supplementary Fig.S2a). Cognitive processes with large positive loadings reflected executive, goal-oriented, and sensorimotor functions (“action,” “skills,” “planning,” “working memory,” and “attention”), whereas emotional, motivational, and interoceptive processes (“sleep,” “anxiety,” “mood,” and “reward anticipation”) showed large negative loadings. The regions with more pronounced cortical thinning in Cluster 1 predominated in the medial and anterior ventral cortices, functionally aligned with the affective terms. Conversely, regions showing relative cortical preservation were localized in the lateral prefrontal and posterior-dorsal cortices, associated with executive and goal-oriented functions (Fig.4a).

**Figure 4.**
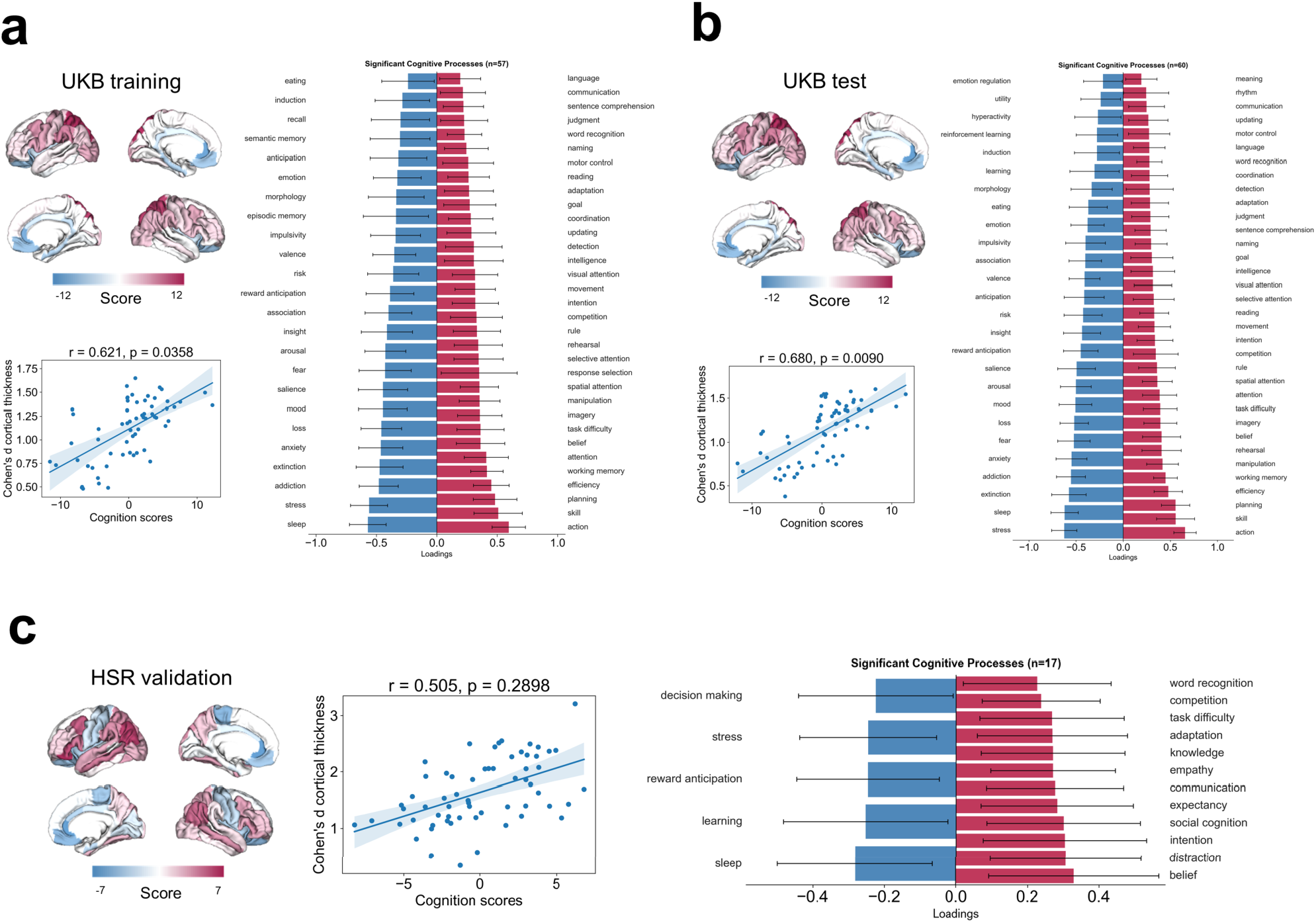
PLS analysis linking cortical thickness patterns to cognitive processes. Each panel (a–c) displays results from a PLS analysis conducted in (a) UKB training set, (b) UKB test set, (c) HSR validation set. Brain maps (top left of each panel): Lateral and medial surface renderings showing regional PLS scores, reflecting how strongly each cortical region contributes to the latent brain-cognition relationship. Warm colors (red/pink) indicate positive scores; cool colors (blue) indicate negative scores. Color scale bars indicate the score range. Scatter plots (bottom left of each panel): Correlation between individual cognition scores (derived from PLS) and Cohen’s d of cortical thickness (y-axis), with Pearson’s r and associated p-value shown. The blue line represents the linear fit with a shaded 95% confidence interval. Each dot represents one brain region. Bar charts (right of each panel): Loadings of significant cognitive process terms (derived from *Neurosynth* meta-analytic maps) on the first PLS component. Blue bars indicate negative loadings; red/pink bars indicate positive loadings. Error bars reflect 95% bootstrap-estimated confidence intervals. UKB, UK Biobank; HSR, San Raffaele Hospital.

These findings were closely replicated in the UKB held-out test set, where a significant LV was found (p_spin_=0.013, r=0.68), also with distance-dependent cross-validation (mean out-of-sample r(60)=0.52, p_spin_=0.001) (Supplementary Fig.S2b). The cognitive term loadings and spatial patterns of PLS brain scores were highly consistent with those from the training set (Fig.4b).

We then examined whether the cluster-specific cortical profiles in HSR map onto the same cognitive processes as in UKB. Although the PLS model did not yield a significant LV (p_spin_=0.28, r=0.51; mean out-of-sample r(60)=0.13, p_spin_=0.22) (Supplementary Fig.S2c), the association with the affective and motivational processes (“sleep”, “stress”, “reward anticipation”, “learning”, and “decision making”) was largely preserved. In contrast, the goal-oriented dimension showed a shift toward higher-order social cognition, with prominent loadings for “belief”, “intention”, “social cognition”, and “empathy”. This partial yet meaningful correspondence extended to the spatial level, where the PLS brain score map retained the overall pattern observed in UKB, though with reduced magnitude and spatial extent (Fig.4c).

### Mapping receptors and transporters to clusters’ cortical profiles

Finally, we asked which neurotransmitter systems underlie the cluster-specific profiles by fitting a multilinear model to predict cluster-specific cortical profiles from PET neurotransmitter maps. The relative contribution of each receptor/transporter to the model fit was estimated via dominance analysis, and robustness was tested through distance-dependent cross-validation (Supplementary Fig.S3).

In UKB, the clusters’ cortical signatures were heavily influenced by receptor distribution (R^2^_adj_=0.73, p_spin_<0.001). The dopamine transporter (DAT) density was the strongest contributor to cluster-specific cortical patterns (15.7%), followed by nicotinic acetylcholine (α4β2) (10.7%), serotonin receptors, and transporters (5HT6:8.10%, 5HTT: 8.60%). The same close fit was found in the UKB hold-out test set (R^2^_adj_=0.76, p_spin_<0.001), where again DAT emerged as a dominant predictor (17.0%).

In HSR, the clusters’ cortical profiles were significantly predicted by receptor and neurotransmitter densities, although with a lower fit compared to UKB (R^2^adj=0.47, p_spin_=0.003). The relative contribution of individual neurotransmitter systems differed from that of UKB, with the histamine receptor H3 emerging as the top contributor (17.2%), followed by 5HT6 (12.0%) and metabotropic glutamate receptor 5 (mGluR5:11.8%), which showed comparable dominance (Fig.5).

**Figure 5.**
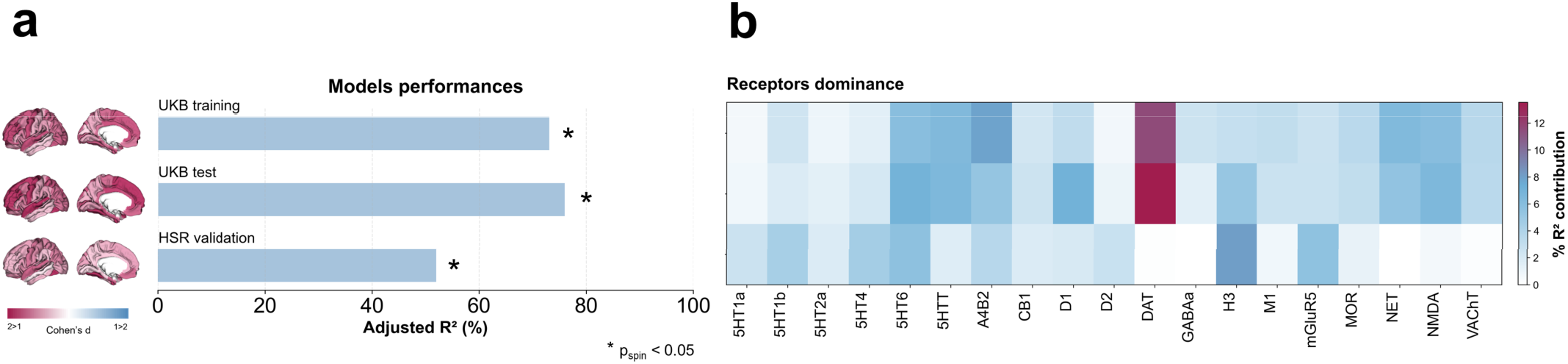
Mapping receptors and transporters di clusters’ cortical profiles. a, For each set (UKB training, UKB test, HSR validation), a multiple linear regression model was fitted between 18 cortical neurotransmitter receptor and transporter density profiles and the cluster-specific cortical pattern (shown as surface plots). Model fits (adjusted R2) are shown in the bar plot. b, Dominance analysis was applied to the independent variables (receptors and transporters) to determine which receptors/transporters were contributing most to the model fit. Percent contribution is shown in the heatmap. UKB, UK Biobank; HSR, San Raffaele Hospital.

## Discussion

In this study, we investigated how regional measures of CT are informative for a brain-based stratification of MDD in a large-scale population cohort, and whether this stratification generalize to an independent cohort of depressed inpatients. Two highly distinguishable clusters emerged in the UKB with 87.5% accuracy, with one cluster characterized by widespread cortical thinning, childhood trauma, and comorbidity of diabetes. This cortical profile generalized with more than 80% accuracy in the HSR cohort, with most patients assigned to the most compromised cluster. By spatially relating the cluster-specific cortical patterns with neurotransmitter receptor profiles and functional activation maps, we provided evidence of the involvement of the dopaminergic system in guiding clusters’ cortical differentiation, alongside a functional dissociation between affective and high-order cognitive processes.

Cluster 1 was characterized by widespread thinning predominantly in the fronto-parieto-temporal cortices, emotional childhood trauma, diabetes comorbidity, and, in the training set only, anhedonia, lethargy, and anxious depression, whereas Cluster 2 showed a relatively preserved cortical structure. Both childhood trauma and diabetes comorbidity have been independently linked to structural brain alterations: childhood trauma, especially the co-occurrence of neglect and abuse, is associated with reduced CT in the temporal and parietal regions^41^, while comorbid diabetes and obesity are similarly related to cortical thinning through inflammatory and mitochondrial mechanisms^42,43^. Importantly, childhood trauma and metabolic dysregulation may not act independently, as the genetic correlation between MDD and body composition has been shown to be specific to individuals with a history of childhood trauma^44^. We hypothesized that the globally reduced CT in Cluster 1 might reflect a cumulative biological burden, in which multiple vulnerabilities converge on brain structure rather than acting through a single etiological pathway.

Going beyond the question of whether these subtypes are cortically and clinically distinct, we then asked what functional processes their cortical signatures reflect. The cortical regions most differentiated between clusters were functionally aligned with meta-analytic maps of affective, motivational, and interoceptive processes, closely mirroring the neurovegetative and somatic symptom dimensions characterizing Cluster 1 (i.e., lethargy, unenthusiasm). Notably, this spatial pattern spanned from unimodal sensorimotor to transmodal regions ^45,46^, suggesting that the clusters’ cortical differences are not spatially random, but rather follow the intrinsic functional organization of the neocortex, consistent with evidence that disorder-related cortical morphology is systematically shaped by molecular and connectome architecture^47^. Bearing in mind the inferential nature of the meta-analytic approach, the functional correspondence with affective, motivational, and interoceptive processes may serve as a bridge between the structural and clinical levels of our analysis, suggesting that the cortical regions most affected in Cluster 1 sustain the affective/cognitive processes that characterize it clinically.

We then turned to the question of whether these signatures also reflect distinct underlying neurotransmitter systems and what this might tell us about the translational relevance of these subtypes. In UKB, we found a strong spatial alignment with the DAT density map. The dopaminergic system has long been implicated in the motivational deficits characteristics of atypical depression^29,48^ and proposed as a key neurobiological correlate of immuno-metabolic depression, characterized by anergy-related depressive symptoms, systemic low-grade inflammation, and metabolic dysregulation^49–51^. Although PET studies of DAT availability in MDD have primally focused on striatal and midbrain regions^52,53^, its expression has been described in prefrontal, anterior cingulate, and motor, and insular cortices^54^, and both chronic stress and inflammation reduce mesocortical dopaminergic integrity through axonal loss and decreased synaptic availability^48,55,56^. Acknowledging the correlational nature of these findings, the convergence across micro- and macro-scale levels indicates a neurobiologically distinct subtype associated with an atypical/metabolic signature. The identified clusters were also structurally and clinically different from HC, with both exhibiting higher depressive symptomatology, childhood trauma, and BMI, and HC cortical profiles falling between the two clusters. This suggests that the stratification captures a neurobiological dimension cutting across diagnostic boundaries rather than a simple pathological/healthy distinction. The relative cortical thickening in Cluster 2 does not necessarily reflect healthier brain functioning: patients were not spared from adversities but lacked the additional co-occurring vulnerabilities of diabetes and childhood abuse that characterized Cluster 1. Cluster 2 may thus represent a distinct subtype where vulnerability still exists, yet without reaching the cumulative burden that drives widespread cortical thinning.

The validation in HSR support the clinical relevance of this stratification. Nearly 80% of HSR patients were assigned to Cluster 1, and, although not surviving correction for multiple comparisons, they were characterized by older age, later onset, greater cardiometabolic comorbidity and treatment-resistance. The spatial association with affective-interoceptive processes was largely preserved, while the emergence of higher-order social cognitive processes in HSR may instead reflect interpersonal and mentalizing difficulties that are more salient in hospitalized patients. The neurochemical mapping, however, substantially diverged from UKB, with histamine receptor H3 making the largest contribution. This may reflect the acute and pharmacologically treated nature of the hospitalized HSR sample, given the established affinity of traditional antidepressants for histaminergic receptors^57^. At the same time, the predominance of H3 may also reflect its broad regulatory role in neurotransmitter release across cortical, striatal, and nigral neural systems by acting as a heteroreceptor on non-histaminergic neurons^58,59^. This is consistent with the comparable contributions of 5HT6 and mGluR5, given that H3 intersects with serotonergic, metabotropic glutamate, and dopaminergic transmission within the thalamocortical and cortico-striatal^58^. Together, these findings may indicate partial continuity with the UKB neurochemical signature, both converging on dopaminergic regulation but through different mechanisms across cohorts. These observations gain further relevance in light of the recent linking of histaminergic gene expression with cortical thinning in MDD and emotion regulation, salience, and stress-related processes^60^. Notably, preclinical evidence suggests that SSRIs require an intact histaminergic system to exert their therapeutic effects and that elevated brain histamine may dampen their serotonergic effects during low-grade inflammation in refractory depression^61,62^. Whether this neurochemical pattern reflects subtype-specific neurobiology, pharmacological confounding, or their interactions remains to be clarified in longitudinal studies with drug-naïve patients.

There are some caveats and limitations that need to be addressed. The moderate sample size of the HSR cohort and the population differences with UKB call for replication in larger demographically and clinically diverse samples. All HSR patients were under pharmacological treatment, making it difficult to disentangle the effects of medication from intrinsic neurobiological correlates. Both PET and *Neurosynth* data were acquired from different samples and protocols. Although this was partially mitigated by focusing on relative spatial topography rather than absolute values^18^, residual influences cannot be excluded. Similar considerations apply to scanner differences between UKB and HSR, which we addressed by preprocessing HSR data with the same UKB pipeline^63^. While the cortical mappings were validated through cross-validation and spatial autocorrelation-preserving null models^45^, their correlational nature warrants caution in drawing direct biological inferences. Finally, analyses were restricted to CT, and future work incorporating subcortical structures may provide a more comprehensive neurobiological characterization.

Overall, this work provides a replicable computational framework for brain-based subtyping of depression, mapping structural and clinical profiles across neurochemical and affective/cognitive dimensions to reveal how subtypes are anchored to multiscale complexity.

## Acknowledgements

The current study was supported by the Italian Ministry of Health, GR-2019-12370616. The presented research has been conducted using the UK Biobank Resource under Application Number 56514 “Stratification of health outcomes in mood disorders”.

## Data availability

Data used to perform the analyses can be found at https://ordr.hsr.it/preview/w67mxvnsy8?a=17d38898-0b42-473a-8fae-ecb8ee357819. Volumetric PET images for neurotransmitter receptors and transporters are included in *neuromaps* (https://github.com/netneurolab/neuromaps). Neurosynth data are available at https://neurosynth.org/.

## Code availability

All code used to perform the analyses can be found at https://github.com/fede-colombo/colombo_ukb_clustering_validation.

## Materials and Methods

### Participants

The UKB is a population-based cohort of approximately 500,000 participants from the United Kingdom. It collects extensive longitudinal data encompassing environmental factors, lifestyle habits, physical activity, genetics, multimodal imaging, diverse types of biomarkers, and health-related information 64. In this study we included individuals of 18-65 years old who showed proxies of MDD, according to at least one of the following criteria: i) diagnosis of a depressive disorder determined from primary care records, using previously described diagnostic codes 65; ii) International Statistical Classification of Diseases (ICD-10) codes for major depressive disorder (F32-F33). Exclusion criteria included any diagnosis of bipolar, psychotic, or substance use disorder identified through primary care records or hospital ICD-10 diagnostic codes. To identify potentially currently depressed individuals at the time of scanning, we selected individuals who scored > 0 on the “Frequency of depressed mood in the last two weeks” item (Data-Field 2050, Instance 2, corresponding to the imaging visit) from the RDS-4 scale. Among them, we selected individuals based on the presence of neuroimaging data, resulting in a final sample of 1,531 participants. To characterize clusters against a sample of non-affected individuals, we extracted HC aged 18-65 years from the UKB who satisfied the following criteria: i) no history of mental illness (based on ICD-10 codes or primary care records) and ii) a score of 0 on the “Frequency of depressed mood in the last two weeks” item (Data-Field 2050, Instance 2) from the RDS-4 (N=954). To ensure comparability between MDD cases and HC, we implemented a greedy matching algorithm to balance groups on age and sex. MDD cases were kept fixed, while the control group sample size was iteratively reduced until statistical balance was achieved (α=0.05) (see Supplementary Methods S1 for additional details). After matching, the final control group sample size was N=827. A complete summary of the subsets of matched HC is presented in Supplementary Table S11. The current study was performed under UKB study ID 56514.

The independent clinical cohort consisted of 144 currently depressed MDD inpatients who were consecutively admitted to the Mood Disorder Unit of IRCCS San Raffaele Hospital in Milan, Italy. Participants were included if they met the Diagnostic and Statistical Manual of Mental Disorders (DSM-5) diagnostic criteria for MDD, as diagnosed by trained psychiatrists, and were aged 18-65 years. Exclusion criteria included psychotic symptoms, other psychiatric comorbidities, intellectual disability, pregnancy, significant medical or neurological conditions, and history of substance abuse. The presence of depressive symptoms at the time of MRI scanning was assessed using the Hamilton Depression Rating Scale (HDRS-21)66. All participants provided written informed consent after receiving a comprehensive description of the study. The local ethics committee approved the study protocol.

### Sociodemographic measures and assessment of clinical outcomes

Outcomes of interest to profile clusters in both the UKB and HSR included sociodemographic variables (i.e., age, sex, education); outcomes related to depression phenotype (i.e., depressive symptomatology, number of depressive episodes, age of onset, treatment resistance, depression with anxious or atypical features); history of childhood trauma; BMI; diabetes; and cardiometabolic comorbidity. A complete list of the selected outcomes in UKB and the corresponding recoding in HSR is provided in Supplementary Tables S12-S13.

### MRI data acquisition

In the UKB cohort, neuroimaging data were acquired using 3T Siemens Skyra scanners at three sites across the United Kingdom. Details about the imaging protocol are reported online at https://biobank.ctsu.ox.ac.uk/crystal/refer.cgi?id=2367 and published in previous works ^67^.

In the HSR cohort, MRI images were acquired using two different 3.0 Tesla scanners. Acquisition of 52 subjects was performed with Gyroscan Intera, Philips, Netherlands employing an 8 channels SENSE head coil (T1-weighted MPRAGE sequences: TR 25.00 ms, TE 4.6 ms, field of view FOV=230 mm,91 matrix=256×256, in-plane resolution 0.9×0.9 mm, yielding 220 transversal slices with a thickness of 0.8 mm). The acquisition of 92 subjects was performed with an Ingenia CX, Philips, The Netherlands using a 32-channel sensitivity encoding SENSE head coil (T1-weighted MPRAGE sequence: TR 8.00 ms, TE 3.7 ms, field of view FOV = 256 mm, matrix = 256 x 256, in-plane resolution 1 x 1 mm, yielding 182 transversal slices with a thickness of 1 mm).

### MRI preprocessing and feature extraction

In UKB, CT imaging-derived phenotypes for each region of the Desikan-Killiany-Tourville (DKT) atlas were extracted (Data-Fields 27174-27296). Data were preprocessed using FreeSurfer 6.0.0, and quality control was performed within the UKB initiative, as previously described^63^. To ensure cross-cohort comparability, T1-weighted images in HSR were preprocessed and checked for quality control according to the UKB pipeline using FreeSurfer 6.0.0, and region-based CT values were extracted based on the DKT atlas. In HSR, to control for potential confounding effects of between-scanner variability, region-based CT values were harmonized using ComBat (*neuroCombat* Python package), a well-established method that uses an empirical Bayesian approach to remove batch effects on neuroimaging data^68^. Age and sex were entered as biological covariates to be preserved from the removal of scanner effects. In UKB, no harmonization across acquisition sites was performed, given that UKB uses identical scanner hardware, software, and protocols across all imaging centres^63^, with empirical validation confirming excellent between-site reliability for morphometric measures^69^. Additional details on the MRI data preprocessing and feature extraction are reported in Supplementary Methods S2.

### Unsupervised machine learning analysis

Unsupervised brain-based clustering was performed using the *NeuReval* Python package (https://github.com/fede-colombo/NeuReval) to perform stability-based relative clustering validation. The mean CT for each region from the DKT atlas was used as input features, considering KMeans as the clustering algorithm due to its computational efficiency and wide adoption in neuroimaging-based stratification studies^70^, and support vector machine (SVM) as the classifier given its robustness in high-dimensional neuroimaging data^71^. All input features were standardized by removing the mean and scaling to the unit variance. The confounding effects of age and sex were removed using linear regression. In addition, age-by-sex interaction and the square of age were considered confounding variables to account for potential non-linear effects of age ^72^. All these steps were fitted to the UKB training set and then applied to the UKB held-out test set. Within the UKB training set, an internal 2-fold cross-validation scheme was employed and repeated 100 times to ensure the robustness of the identified clustering solutions ^73^. A grid search procedure was performed within the internal cross-validation to tune the hyperparameters for both the clustering and classifier algorithms, using the minimization of the normalized stability as the model selection criterion. These included the soft-margin C (range: 0.01, 0.1, 1, 10, 100, 1000) and the type of kernel (i.e., linear or radial basis function) for SVM. The entire process was iterated over a number of clusters from 2 to 4, a range consistent with the neurobiologically meaningful subtypes reported in the depression literature^28^. The clustering solution that achieved the minimum normalized stability throughout the cross-validation was selected. The optimal number of clusters and the SVM hyper-parameters fitted on the full training set were then applied to the UKB held-out test set to assess generalization. The generalization accuracy of the UKB held-out test set was then computed by comparing the classifier’s predicted labels with the actual clustering labels identified in the held-out set.

In addition to the normalized stability, the silhouette score and Davies-Bouldin index were computed for each stratification model. Finally, to verify whether the identified clustering solutions reflect evidence consistent with the cluster structure, the *sigclust* method (R library) with 10,000 Monte Carlo simulations was applied to each feature set, considering the same standardization and correction for nuisance covariates implemented in *NeuReval* ^74^.

### Characterizations of clusters for input and outcome variables

In the case of significant stratification, clusters were characterized for regional CT values and outcome variables not used as input features (i.e., sociodemographic and depression-related variables, childhood trauma, diabetes, and cardiometabolic comorbidity). Clusters’ characterization was conducted separately and independently within the UKB training and held-out test sets, with consistent results across both sets considered as evidence of robustness.

Between-cluster differences in regional CT were assessed using two-sample t-tests or Wilcoxon-Mann-Whitney tests, depending on normality assumptions verified via the Shapiro-Wilk test, with Cohen’s d as the effect size measure. Differences between clusters and HC for both regional and global CT, as well as for continuous outcome variables, were evaluated using ANOVA or Kruskal-Wallis tests, with pairwise post-hoc comparisons in case of significant group effects and eta-squared as the effect size measure. Categorical outcome variables were compared using χ^2^ tests or Fisher’s exact tests, with Cramér’s V as the effect size measure. Individuals with missing data were excluded from the analyses. Multiple comparisons were corrected using the Benjamini-Hochberg False Discovery Rate (FDR) procedure, with q<0.05 as the significance threshold.

### Mapping meta-analytic cognitive functions to clusters’ cortical profiles

Meta-analytic functional activation maps were derived from *Neurosynth*, a meta-analytic database comprising more than 15,000 functional MRI studies (www.neurosynth.org). For each brain voxel, the extracted value reflects the probability that the voxel is activated during tasks associated with a specific meta-analytic term. Consistent with previous studies^18^, this analysis focused on 123 cognitive process terms derived from the Cognitive Atlas (www.cognitiveatlas.org), a public ontology of cognitive science that provides a comprehensive list of neurocognitive processes^75^. Data derived from *Neurosynth* were parcellated using the DKT atlas and subsequently z-scored. The probabilistic measure from *Neurosynth* serves as a quantitative indicator of the relationship between regional activity fluctuations and the psychological processes. The complete list of cognitive processes is provided in Supplementary Table S14.

To investigate whether the spatial distribution of cluster-specific cortical differences aligns with meta-analytic maps of cognitive and functional processes, we performed a spatial association analysis between cluster-derived cortical effect size maps and *Neurosynth* functional activation maps. We applied PLS independently in both the UKB training and held-out test sets, with convergent results across sets considered as evidence of robustness of the identified mapping patterns. PLS is a data-driven multivariate technique that identifies latent variables capturing the maximum covariance between two sets of variables (in this case, CT Cohen’s d values (X) and functional activations (Y) for each region of DKT atlas)^76^. This yields a set of *Neurosynth* terms weights that can be aggregated across brain regions and expressed as cognitive scores, producing a summary map of the functional terms most spatially associated with the observed cortical pattern. Statistical significance of LV was tested through 10,000 permutations spin-tests to control for spatial autocorrelations^77^. Specifically, the spatial coordinates for each parcel were determined through a spherical projection of the *fsaverage* surface, locating the vertex most proximal to the center of the mass of each parcel. Random rotations were then applied to these coordinates, and each original parcel was reassigned the value of its closest rotated equivalent, with this process repeated 10,000 times. In cases where the nearest match of a parcel was the medial wall, the value was substituted with that from the next closest parcel. To further assess the robustness of the identified spatial correspondence, a distance-dependent cross-validation was implemented. Briefly, a PLS model was fitted on the training set (75%) and derived weights were applied on the test set (25%) to generate predicted cognitive and cortical scores. The statistical significance of the resulting out-of-sample correlations was evaluated against a permutation-based null model, generated by repeating the cross-validation procedure on 1,000 spatially constrained permutations of the parcellated *Neurosynth* functional terms matrix (1,000 iterations)^18^. More details about PLS analysis are provided in Supplementary Methods S3.

### Mapping neurotrasmitters receptors and transporters to clusters’ cortical profiles

To investigate the spatial correspondence between cluster-specific cortical patterns and neurotransmitter receptors and transporter maps, we used data compiled by Hansen et al.^18^, accessed through the *neuromaps* Python package. *Neuromaps* is a comprehensive toolbox providing curated brain maps and analytical tools designed to compare different brain maps. In this study, we considered atlas data from more than 1,200 individuals for 19 receptors and transporters, encompassing nine neurotransmitter systems. These included: dopamine (D1, D2, DAT)^78–81^, norepinephrine (NET)^82^, serotonin (5-HT1A, 5-HT1B, HT2A, 5-HT4, 5-HT6,5-HTT)^83–86^, acetylcholine (α4β2, M1, VAChT)^87–90^, glutamate (mGluR5, NMDA)^91–93^, GABAergic (GABAA)^94^, histamine (H3)^84^, cannabinoid (CB1)^95^, and opioid (MOR)^96^. In this study, All PET images were registered to the MNI-ICBM 152 non-linear 2009 (version c, asymmetric) template and parcellated into 62 regions based on the DKT atlas. When multiple mean images existed for the same tracer (i.e., 5-HT1B, D2, mGluR5, and VAChT), data were merged through weighted average calculations, as previously described^18^. The parcellated PET data were then z-scored and concatenated into a region-by-receptor matrix for further analyses. A complete list of neurotransmitters and transporters are reported in Supplementary Table S15.

To explore putative neurochemical correlates of the cluster-specific cortical patterns, we performed a spatial association analysis between cluster-derived cortical effect size maps and normative neurotransmitter receptor and transporter density maps. We note that this approach does not provide a direct measure of neurochemical alterations in our patients. Rather, it tests whether the spatial distribution of CT differences between clusters (expressed as Cohen’s d values for each brain region) aligns with the regional distribution of specific neurotransmitter systems, as captured by normative PET-derived maps parcellated to the DKT atlas. To this end, multilinear regression models were independently fitted in the UKB training and held-out test sets, with cluster-specific Cohen’s d maps as the dependent variable and normative receptor and transporter density maps as predictors.

To estimate the relative contribution of each neurotransmitter system to the overall spatial alignment, we applied dominance analysis ^97^. This approach quantifies the individual contribution of each predictor by fitting the regression model across all possible subsets of input variables (2^p^-1 submodels for *p* predictors), and computing the mean relative increase in R^2^ attributable to each predictor across all configurations. Since the sum of all dominance values equals the total adjusted R^2^ of the complete model, total dominance represents an interpretable metric for assessing the individual contribution of each predictor ^97^. Statistical significance was assessed using 10,000 permutation spin-tests, as described above. To ensure the robustness of each multilinear model and minimize the effect of spatial autocorrelations, a distance-dependent cross-validation approach was implemented ^45^. Briefly, for each brain region serving as a source node, the 75% nearest regions were selected as the training set, and the remaining 25% were selected as the test set. This yielded 62 iterations for the DKT atlas, corresponding to the total number of parcels.

### External validation in inpatient clinical cohort

To externally validate the clusters identified in the UKB cohort, we utilized the HSR as an independent external cohort. Specifically, KMeans with the optimal number of clusters identified in UKB was applied to the HSR cohort, and the SVM model trained in UKB was applied to predict cluster labels. For the stratification model in the UKB, Silhouette and Davies-Bouldin scores were computed, and the significance of the clustering solution was tested using 10,000 permutation tests. Clusters were then profiled for demographic and clinical outcome variables.

Given that population heterogeneity is known to affect the neurobiological interpretation of brain-derived patterns across cohorts^98^, we applied a propensity score full matching in HSR to derive harmonized cortical effect size maps, which were then used for the functional and neurotransmitter mapping. Propensity scores represent the probability of belonging to a given cohort (UKB vs HSR) conditional on observed characteristics significantly different between cohorts (in this case, age, BMI, and sex). Rather than adjusting for each covariate separately, propensity scores model multiple confounding variables into a single balancing score, providing a unified approach to account for between-cohort differences^98,99^. Full matching was performed using the *MatchIt* R package, retaining all available participants in HSR^100^. Each participant was assigned a matching weight reflecting their contribution to the matched pseudo-population (i.e., how much they should count in the analysis to make the two cohorts comparable). Matching weights were then used to obtain harmonized estimates of between-cluster differences in CT. For each cortical region, cluster differences were tested using weighted t-tests, with matching weights added as covariate to account for the pseudo-population structure. Effect sizes were quantified as weighted Cohen’s d, representing the magnitude of confound-adjusted between-cluster differences in CT at each region. The resulting maps of weighted Cohen’s d values (one value per cortical region) were then used as clusters’ cortical profile maps, serving as input for the cognitive function mapping (*Neurosynth*) and neurotransmitter receptor/transporter mapping analyses, following the same pipeline applied in UKB. More details on propensity score full matching are reported in Supplementary Methods S4.

