## Supplementary for "Mapping generalizable brain-based depression subtypes across clinical, cognitive, and neurotransmitter dimensions"

***Supplementary Materials***

### Methods S1. Greedy matching of controls to cases in UKB

To reduce systematic differences between cases and controls on age, sex, and BMI, a greedy iterative reduction algorithm was applied^1^. All subjects with missing values in any of the three matching variables were excluded prior to matching. At each attempt, a random subset of the available control pool was drawn without replacement, with the initial subset size sampled uniformly from 70–100% of the total control pool. Balance between the drawn control subset and the fixed case group was evaluated using Welch's two-sample t-test for age and BMI and either Pearson's chi-squared test or Fisher's exact test for sex (the latter applied when any expected cell count fell below 5). A subset was considered adequately matched when all three p-values exceeded α = 0.05. If the initial subset failed to meet this criterion, an iterative removal procedure was initiated. At each step, a random sample of up to 60 candidate controls was drawn from the current subset, and the effect of removing each candidate was evaluated by recomputing all three p-values. The candidate whose removal produced the greatest improvement in the minimum p-value across matching variables (i.e., the worst-performing test) was permanently removed. In cases of ties in this criterion, the removal that maximized the sum of all three p-values was preferred. This process continued until all three p-values exceeded the threshold, the subset fell below the minimum allowable size (n = 10), or a maximum of 2,000 removal steps was reached without achieving balance. Up to 30 independent attempts were performed, each initialized with a different random starting subset. Each attempt that yielded a balanced subset was retained, and up to 15 unique valid subsets were stored. Subsets were ranked primarily by the number of retained controls (larger preferred) and secondarily by the minimum p-value across matching variables (higher preferred), ensuring that the best-matched subset maximized control sample size while maintaining statistical equivalence. Duplicate subsets (identical sets of control indices) were deduplicated prior to final ranking. The best ranking subset was selected for all subsequent analyses.

### Methods S2. Details of MRI preprocessing in UKB

T1-weighted images were acquired with a three-dimensional magnetization-prepared rapid acquisition 21 with gradient echo sequence (MPRAGE) (resolution: 1.0 x 1.0 x 1.0 mm; field-of-view matrix: 208 x 256 x 256; 22 inversion time, 880 msec; repetition time, 2000 msec). The T1-weighted pipeline encompassed: i) gradient distortion correction (GDC); ii) field-of-view reduction, to reduce the amount of non-brain tissue; iii) linear and non-linear registration to the MNI152 space, using FMRIB's Linear Image Registration Tool (FLIRT)^2^ and FMRIB's Nonlinear Image Registration Tool (FNIRT16)^3^. The 1 mm resolution version of MNI152 template was used as reference space; iv) brain extraction using Brain Extraction Tool (BET)^4^. Cortical thickness obtained using Freesurfer according to the Desikan-Killiany atlas were employed (UKB Data fields within the Category 196, [Freesurfer DKT](https://biobank.ndph.ox.ac.uk/showcase/label.cgi?id=196)).

### Methods S3. Partial Least Squares analysis

Partial Least Squares (PLS) analysis is a data-driven multivariate technique that identifies latent variables (LVs) capturing the maximum covariance between two sets of variables. The cross-covariance matrix between z-scored Cohen’s d CT values (X) and functional activations of cognitive processes (Y) for each DKT atlas region was computed as:

*R = Xᵀ Y*

And decomposed via singular value decomposition (SDV):

*R = U Σ Vᵀ*

where *U* and *V* contain the left and right singular vectors (imaging and cognitive weights, respectively), and Σ is a diagonal matrix of singular values. Each pair of singular vectors defines a latent variable (LV), ordered by the amount of cross-covariance explained. The left singular vectors (i.e. the columns of U) represent the degree to which each regional cluster-specific cortical pattern contributes to the latent variable and demonstrates the extracted association between clusters’ cortical profiles and cognitive activation (“CT weights”). The right singular vectors (i.e. the columns of V) represent the degree to which the cognitive terms contribute to the same latent variable (“cognitive weights”). Positively weighed CT profiles covary with positively weighed cognitive terms, and negatively weighed CT profiles covary with negatively weighed cognitive terms. The brain scores and cognitive scores for each subject were computed by projecting the data onto the singular vectors, defined as:

CT scores = *X U*

Cognitive scores = *Y V*

Positively scored brain regions are regions that demonstrate the covariance between expression of positively weighted CT profiles and activation of positively weighted cognitive terms (and vice versa for negatively scored brain regions). The robustness of the PLS model was assessed by cross-validating the correlation between CT scores and term scores. Since in this case observations are brain areas and therefore non-independent, cross-validation was designed such that the training and testing set were made of spatially distant brain regions. To achieve this, the training set was composed of a random source node and the 75% of brain regions closest in Euclidean distance, while the remaining 25% of brain regions were included in the test set. CT scores and cognitive scores were computed in the training set, as well as the correlation between the two (Corr(X_train_U_train_, Y_train_V_train_)). The test set was projected onto the training-derived singular vector weights to obtain predicted CT and cognitive scores, and the correlation between predicted scores was calculated (Corr(X_test_U_train_, Y_test_V_train_)). This procedure was repeated for each brain region as source mode (62 times), yielding a distribution of score correlations for the training and testing sets.

### Methods S4. Propensity score full matching

Full matching was performed using the MatchIt R package^5^, retaining all HSR participants and forming matched sets of individuals from both cohorts with similar propensity scores. Propensity scores were estimated via logistic regression, with cohort membership as the dependent variable and age and BMI as covariates. An exact matching constraint on sex was imposed, such that matched sets included only participants of the same sex^6^. Each participant was assigned a matching weight reflecting their contribution to the matched pseudo-population, ensuring covariate balance between groups. Balance was assessed using standardized mean differences (SMD) before and after matching, with SMD < 0.1 considered indicative of adequate balance^7^. To test the significance of between-cluster differences for each cortical region, weighted t-tests were conducted using weighted least squares (WLS) regression, with cluster membership as the independent variable and matching weights applied to account for the pseudo-population structure. Cluster-robust standard errors were estimated using matched set identifiers as clustering units, to account for the non-independence of observations within matched sets^8^. Multiple comparisons were corrected using the FDR procedure (q=0.05). Effect sizes were quantified using weighted Cohen's d, derived from weighted group means and standard deviations, with the pooled standard deviation estimated as:

$${SD}_{pooled}=\sqrt{\frac{\left( n_{1}-1 \right)s_{1}^{2}+(n_{2}-1)s_{2}^{2}}{n_{1}+n_{2}-2}}$$

where n₁ and n₂ correspond to the sum of weights in each group, and s₁ and s₂ are the respective weighted standard deviations^7^. The derived weighted Cohen's d values were then used as input to meta-analytic cognitive function mapping and neurotransmitter receptor and transporter mapping, following the same analytical pipeline described for the UKB cohort.

### Figure S1. Differences in global cortical thickness between clusters and healthy controls.

Results are separately reported for UKB training (a) and test set (b). Asterisks indicate statistically significant pairwise group comparisons that survive false discovery rate (FDR) q < 0.05. UKB, UK Biobank

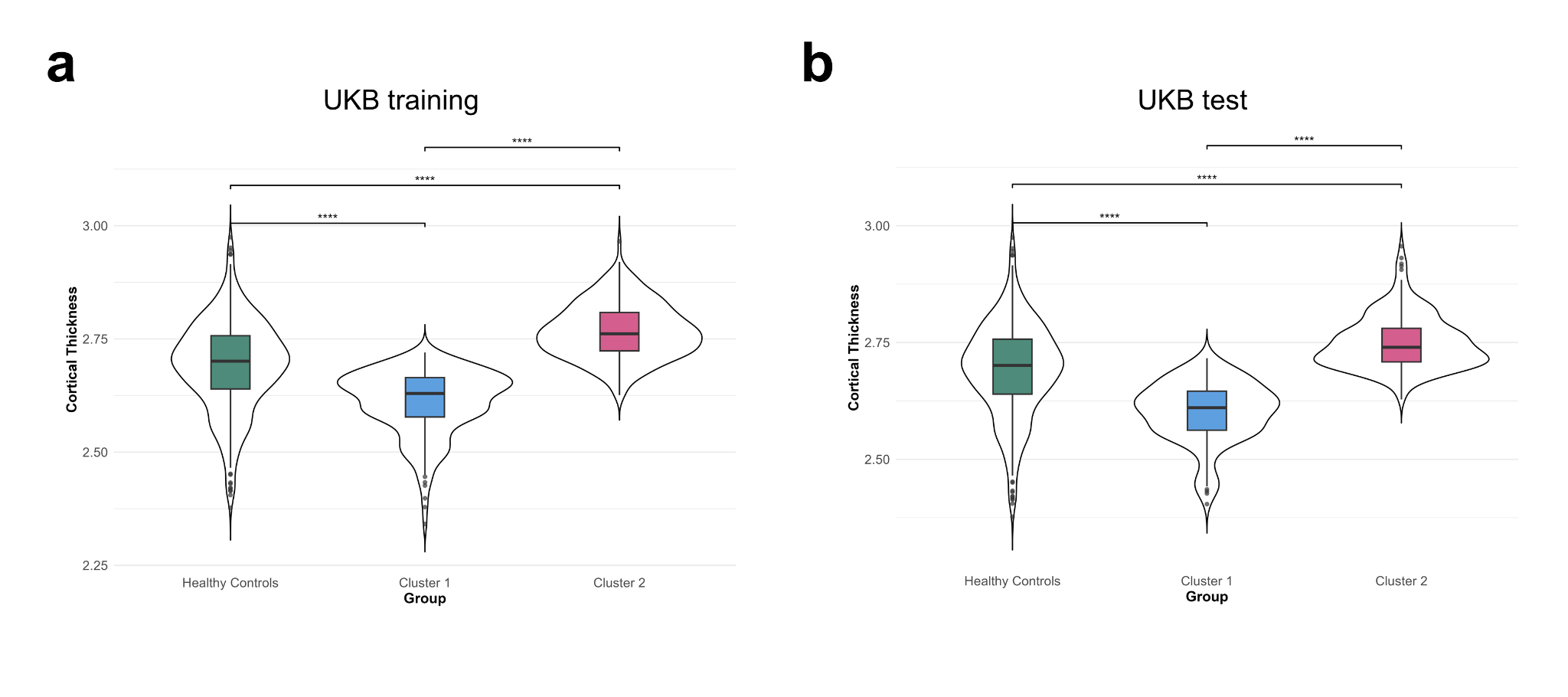

### Figure S2. Cross-validating models to map cluster-specific cortical profiles onto cognitive/affective processes.

For each set (UKB training, UKB test, HSR validation), partial least squares (PLS) models mapping cluster-specific cortical profiles onto Neurosynth-derived cognitive functional activations were cross-validated using a distance-dependent method. For each iteration, the 25% of regions closest to a source region were held out as the test set, while the remaining 75% of regions served as the training set. This procedure was repeated for each of the 62 brain regions as the source region. Model performance was assessed by correlating the predicted cortical pattern with the empirical cortical pattern in the test set. UKB, UK Biobank; HSR, San Raffaele Hospital.

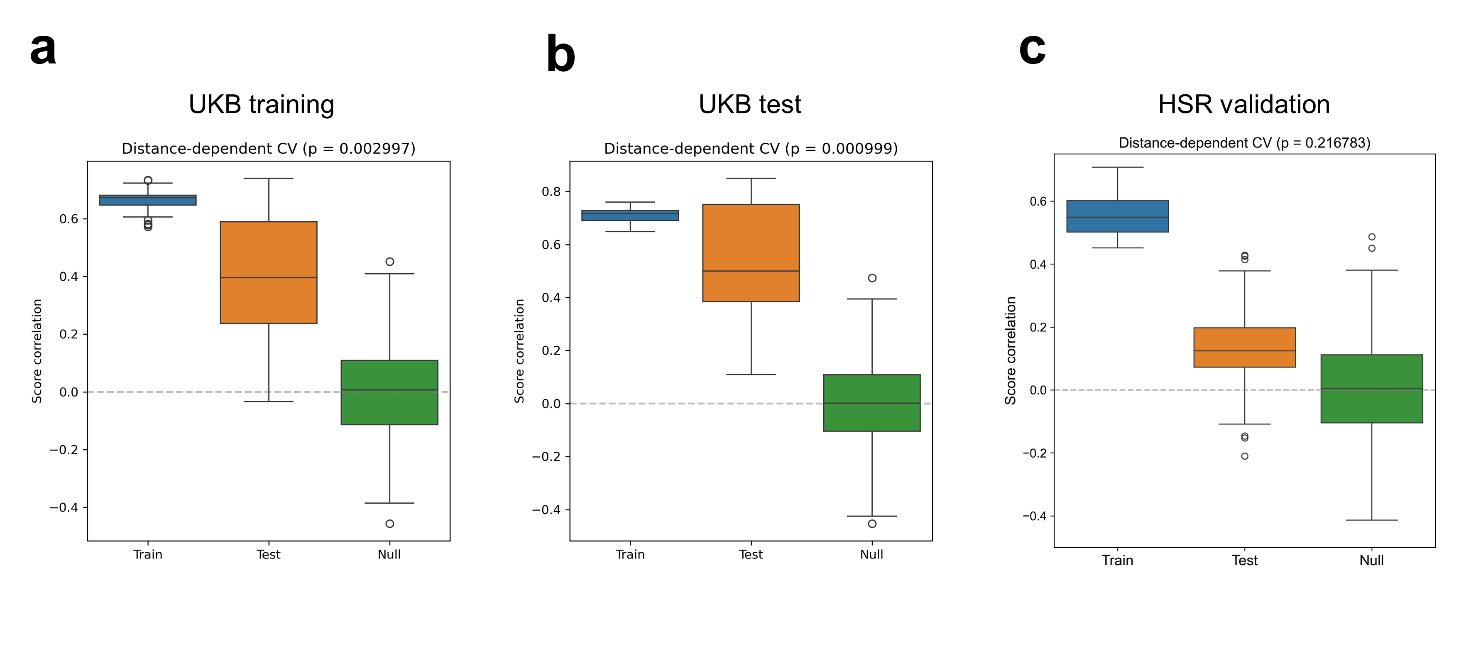

### Figure S3. Cross-validating models that predict cluster-specific cortical profiles from receptor/transporter densities.

For each set (UKB training, UKB test, HSR validation), multilinear models between receptor/transporter densities and cluster-specific cortical profiles were cross-validated using a distance-dependent method. For each iteration, the 25% of regions closest to a source region were held out as the test set, while the remaining 75% of regions served as the training set. This procedure was repeated for each of the 62 brain regions as the source region. Model performance was assessed by correlating the predicted cortical pattern with the empirical cortical pattern in the test set. UKB, UK Biobank; HSR, San Raffaele Hospital

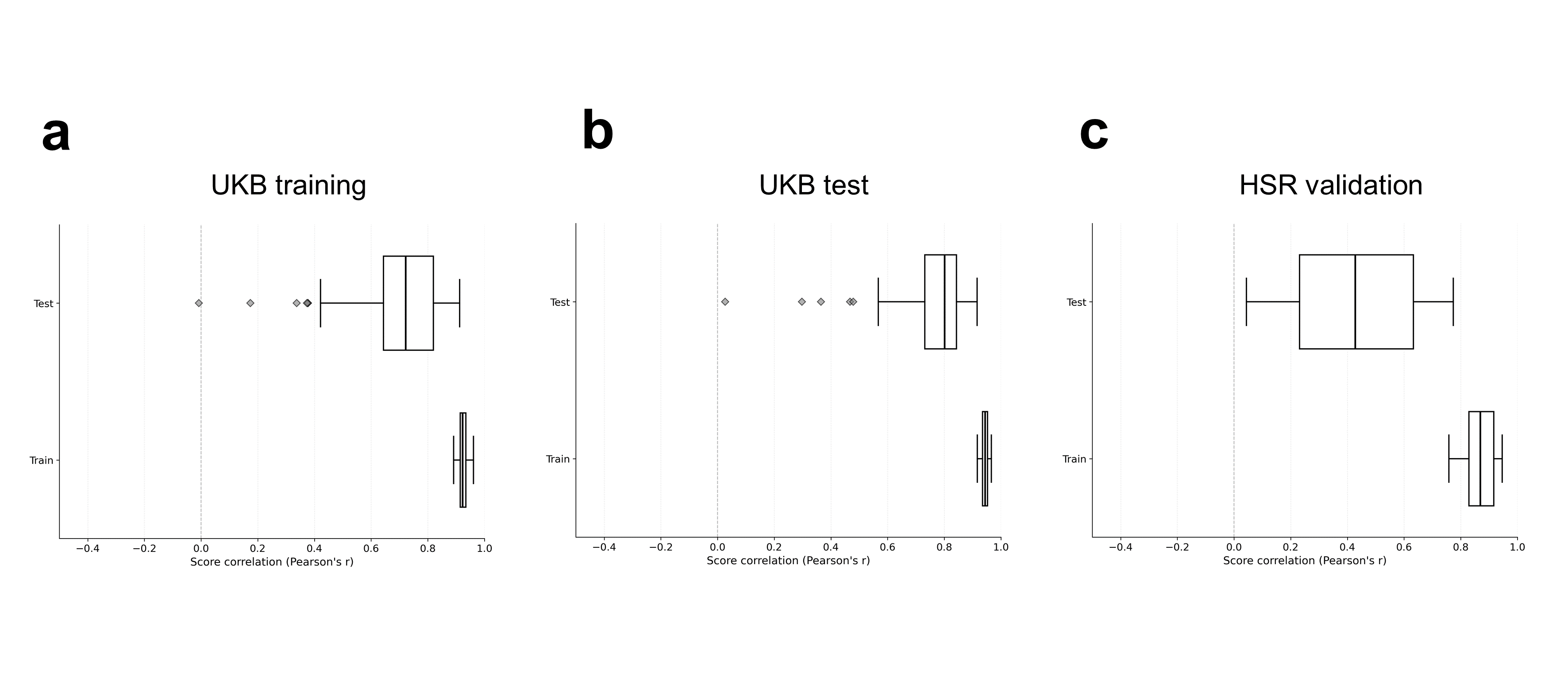
