## Supplementary Table for "Mapping generalizable brain-based depression subtypes across clinical, cognitive, and neurotransmitter dimensions"

***Supplementary Tables***

Table S1. Comparisons between UKB and HSR cohorts for common outcome variables

Table S2. Comparisons between UKB train and test sets for outcome variables

Table S3. Comparisons on outcome variables between healthy controls and MDD patients in UKB cohort

Table S4. Summary of stratification models' performances

Table S5. Comparisons on cortical thickness regional values (DKT atlas) between healthy controls and UKB clusters from the training set

Table S6. Comparisons on cortical thickness regional values (DKT atlas) between healthy controls and UKB clusters from the test set

Table S7. Comparisons on outcome variables between healthy controls and UKB clusters from the training set

Table S8. Comparisons on outcome variables between healthy controls and UKB clusters from the test set

Table S9. Weighted comparisons on cortical thickness regional values (DKT atlas) between clusters from HSR validation cohort

Table S10. Comparisons on outcome variables between clusters from the HSR validation cohort

Table S11. Summary of subsets of matched healhty controls derived from greedy matching algorithm

Table S12. List of outcome variables included in UKB and HSR cohorts

Table S13. List of drugs included in antidepressant medications in HSR cohort

Table S14. Neurosynth terms

Table S15. Neurotransmitter receptors and transporters used to build the receptor density matrix.

**Table S1. Comparisons between UKB and HSR cohorts for common outcome variables**

| **Variable** | **UKB (N=1,531)** | **HSR (N=144)** | **ES** | **p** | **q** |
| --- | --- | --- | --- | --- | --- |
| Age (years) | 58.00 (53.00–61.00) | 53.00 (44.75–58.25) | d = 1.179 | <0.001 | <0.001 |
| Sex (% male) | 485/1531 (31.7%) | 55/144 (38.2%) | d = 0.037 | 0.132 | 0.198 |
| BMI | 27.10 (23.80–30.90) | 24.47 (21.67–28.40) | d = 0.487 | <0.001 | <0.001 |
| Age of onset (years) | 41.00 (31.00–49.00) | 31.00 (25.00–40.00) | d = 0.511 | <0.001 | <0.001 |
| Number of depressive episodes | 4.00 (2.00–8.00) | 3.50 (2.00–6.00) | d = 0.195 | 0.619 | 0.619 |
| Diabetes / cardiometabolic comorbidity | 106/1528 (6.9%) | 13/144 (9.0%) | d = 0.019 | 0.445 | 0.501 |
| Depression with anxious features | 497/1463 (34.0%) | 43/144 (29.9%) | d = 0.023 | 0.366 | 0.471 |
| Depression with atypical features | 151/1445 (10.4%) | 3/140 (2.1%) | d = 0.076 | 0.003 | 0.005 |
| Treatment resistant depression | 64/565 (11.3%) | 53/140 (37.9%) | d = 0.28 | <0.001 | <0.001 |
| Abbreviations: UKB, UK Biobank; HSR, San Raffaele Hospital; ES, Effect Size; BMI, Body Mass Index. | | | | | |

**Table S2. Comparisons between UKB train and test sets for outcome variables**

| **Variable** | **UKB train set (N=842)** | **UKB test set (N=689)** | **ES** | **p** | **q** |
| --- | --- | --- | --- | --- | --- |
| Age (years) | 58.00 (53.25–61.00) | 57.00 (53.00–61.00) | d = 0.037 | 0.522 | 0.849 |
| Sex (% males) | 267/842 (31.7%) | 218/689 (31.6%) | V = 0.000 | 1 | 1 |
| BMI | 27.00 (23.80–30.60) | 27.20 (23.92–31.10) | d = -0.049 | 0.455 | 0.849 |
| Education | 6.00 (3.00–6.00) | 5.00 (3.00–6.00) | d = 0.015 | 0.717 | 0.849 |
| Age of onset (years) | 41.00 (31.00–49.25) | 40.00 (30.00–48.00) | d = 0.059 | 0.369 | 0.849 |
| Number of depressive episodes | 4.00 (2.00–8.00) | 4.00 (2.00–7.00) | d = 0.051 | 0.41 | 0.849 |
| Antidepressant treatment | 261/607 (43.0%) | 206/501 (41.1%) | V = 0.017 | 0.569 | 0.849 |
| Diabetes | 71/841 (8.4%) | 35/687 (5.1%) | V = 0.063 | 0.014 | 0.308 |
| Depression with anxious features | 275/811 (33.9%) | 222/652 (34.0%) | V = 0.000 | 0.999 | 1 |
| Depression with atypical features | 86/799 (10.8%) | 65/646 (10.1%) | V = 0.009 | 0.729 | 0.849 |
| Treatment resistant depression | 34/318 (10.7%) | 30/247 (12.1%) | V = 0.017 | 0.684 | 0.849 |
| RDS-4 Total score | 4.00 (3.00–6.00) | 4.00 (3.00–6.00) | d = -0.059 | 0.502 | 0.849 |
| RSD-4 Depressed mood | 1: 640 (76.0%); 2: 108 (12.8%); 3: 94 (11.2%) | 1: 528 (76.6%); 2: 78 (11.3%); 3: 83 (12.0%) | d = -0.004 | 0.877 | 0.965 |
| RSD-4 Lethargy | 0.0: 300 (36.3%); 1.0: 394 (47.7%); 2.0: 69 (8.4%); 3.0: 63 (7.6%) | 0.0: 225 (33.3%); 1.0: 333 (49.3%); 2.0: 56 (8.3%); 3.0: 62 (9.2%) | d = -0.070 | 0.188 | 0.849 |
| RSD-4 Tensensess | 0.0: 101 (12.1%); 1.0: 431 (51.7%); 2.0: 125 (15.0%); 3.0: 177 (21.2%) | 0.0: 92 (13.4%); 1.0: 318 (46.3%); 2.0: 121 (17.6%); 3.0: 156 (22.7%) | d = -0.044 | 0.374 | 0.849 |
| RSD-4 Unenthusiasm | 0.0: 216 (26.1%); 1.0: 468 (56.5%); 2.0: 87 (10.5%); 3.0: 58 (7.0%) | 0.0: 178 (26.3%); 1.0: 367 (54.3%); 2.0: 69 (10.2%); 3.0: 62 (9.2%) | d = -0.046 | 0.599 | 0.849 |
| CTS-5 Total score | 2.00 (0.25–5.00) | 2.00 (1.00–5.00) | d = -0.070 | 0.733 | 0.849 |
| CTS-5 Emotional Neglect | 0.0: 271 (34.0%); 1.0: 183 (23.0%); 2.0: 222 (27.9%); 3.0: 86 (10.8%); 4.0: 35 (4.4%) | 0.0: 209 (32.6%); 1.0: 143 (22.3%); 2.0: 188 (29.3%); 3.0: 75 (11.7%); 4.0: 26 (4.1%) | d = -0.032 | 0.496 | 0.849 |
| CTS-5 Emotional Abuse | 0.0: 524 (66.1%); 1.0: 75 (9.5%); 2.0: 121 (15.3%); 3.0: 37 (4.7%); 4.0: 36 (4.5%) | 0.0: 412 (64.1%); 1.0: 68 (10.6%); 2.0: 92 (14.3%); 3.0: 35 (5.4%); 4.0: 36 (5.6%) | d = -0.049 | 0.398 | 0.849 |
| CTS-5 Physical Abuse | 0.0: 555 (69.9%); 1.0: 111 (14.0%); 2.0: 97 (12.2%); 3.0: 19 (2.4%); 4.0: 12 (1.5%) | 0.0: 427 (66.2%); 1.0: 95 (14.7%); 2.0: 97 (15.0%); 3.0: 16 (2.5%); 4.0: 10 (1.6%) | d = -0.074 | 0.122 | 0.849 |
| CTS-5 Sexual Abuse | 0.0: 661 (84.1%); 1.0: 54 (6.9%); 2.0: 52 (6.6%); 3.0: 9 (1.1%); 4.0: 10 (1.3%) | 0.0: 536 (85.4%); 1.0: 34 (5.4%); 2.0: 38 (6.1%); 3.0: 9 (1.4%); 4.0: 11 (1.8%) | d = -0.003 | 0.591 | 0.849 |
| CTS-5 Physical Neglect | 0.0: 653 (82.3%); 1.0: 82 (10.3%); 2.0: 30 (3.8%); 3.0: 11 (1.4%); 4.0: 17 (2.1%) | 0.0: 506 (78.8%); 1.0: 67 (10.4%); 2.0: 40 (6.2%); 3.0: 10 (1.6%); 4.0: 19 (3.0%) | d = -0.104 | 0.069 | 0.759 |
| Abbreviations: UKB, UK Biobank; ES, Effect Size; BMI, Body Mass Index; RSD-4, Recent Depressive Symptoms; CTS-5, Childhood Trauma Screener | | | | | |

**Table S3. Comparisons on outcome variables between healthy controls and MDD patients in UKB cohort**

| **Variable** | **HC (N=827)** | **MDD (N=1531)** | **ES** | **p** | **q** |
| --- | --- | --- | --- | --- | --- |
| Age (years) | 58.00 (55.00–61.00) | 58.00 (53.00–61.00) | d = 0.081 | 0.068 | 0.077 |
| Sex (% males) | 262/827 (31.7%) | 485/1531 (31.7%) | V = 0.000 | 1 | 1 |
| BMI | 25.63 (23.27–28.43) | 27.10 (23.80–30.90) | d = -0.283 | <0.001 | <0.001 |
| Education | 6.00 (4.00–6.00) | 5.00 (3.00–6.00) | d = 0.071 | 0.11 | 0.118 |
| Diabetes | 32/827 (3.9%) | 106/1528 (6.9%) | V = 0.060 | 0.003 | 0.004 |
| RDS-4 Total score | 1.00 (0.00–1.00) | 4.00 (3.00–6.00) | d = -1.769 | <0.001 | <0.001 |
| RSD-4 Depressed mood | 0: 827 (100.0%) | 0: 29 (1.9%); 1: 1151 (75.2%); 2: 175 (11.4%); 3: 176 (11.5%) | d = -2.355 | <0.001 | <0.001 |
| RSD-4 Lethargy | 0.0: 445 (54.7%); 1.0: 311 (38.3%); 2.0: 31 (3.8%); 3.0: 26 (3.2%) | 0.0: 525 (35.0%); 1.0: 727 (48.4%); 2.0: 125 (8.3%); 3.0: 125 (8.3%) | d = -0.421 | <0.001 | <0.001 |
| RSD-4 Tensensess | 0.0: 629 (77.7%); 1.0: 164 (20.2%); 2.0: 11 (1.4%); 3.0: 6 (0.7%) | 0.0: 193 (12.7%); 1.0: 749 (49.2%); 2.0: 246 (16.2%); 3.0: 333 (21.9%) | d = -1.454 | <0.001 | <0.001 |
| RSD-4 Unenthusiasm | 0.0: 768 (93.7%); 1.0: 46 (5.6%); 2.0: 2 (0.2%); 3.0: 4 (0.5%) | 0.0: 394 (26.2%); 1.0: 835 (55.5%); 2.0: 156 (10.4%); 3.0: 120 (8.0%) | d = -1.336 | <0.001 | <0.001 |
| CTS-5 Total score | 1.00 (0.00–3.00) | 2.00 (1.00–5.00) | d = -0.385 | <0.001 | <0.001 |
| CTS-5 Emotional Neglect | 0.0: 423 (51.2%); 1.0: 197 (23.8%); 2.0: 152 (18.4%); 3.0: 41 (5.0%); 4.0: 13 (1.6%) | 0.0: 480 (33.4%); 1.0: 326 (22.7%); 2.0: 410 (28.5%); 3.0: 161 (11.2%); 4.0: 61 (4.2%) | d = -0.437 | <0.001 | <0.001 |
| CTS-5 Emotional Abuse | 0.0: 677 (82.1%); 1.0: 63 (7.6%); 2.0: 56 (6.8%); 3.0: 17 (2.1%); 4.0: 12 (1.5%) | 0.0: 936 (65.2%); 1.0: 143 (10.0%); 2.0: 213 (14.8%); 3.0: 72 (5.0%); 4.0: 72 (5.0%) | d = -0.392 | <0.001 | <0.001 |
| CTS-5 Physical Abuse | 0.0: 617 (74.7%); 1.0: 126 (15.3%); 2.0: 65 (7.9%); 3.0: 8 (1.0%); 4.0: 10 (1.2%) | 0.0: 982 (68.2%); 1.0: 206 (14.3%); 2.0: 194 (13.5%); 3.0: 35 (2.4%); 4.0: 22 (1.5%) | d = -0.183 | <0.001 | <0.001 |
| CTS-5 Sexual Abuse | 0.0: 734 (89.6%); 1.0: 35 (4.3%); 2.0: 35 (4.3%); 3.0: 10 (1.2%); 4.0: 5 (0.6%) | 0.0: 1197 (84.7%); 1.0: 88 (6.2%); 2.0: 90 (6.4%); 3.0: 18 (1.3%); 4.0: 21 (1.5%) | d = -0.137 | <0.001 | 0.001 |
| CTS-5 Physical Neglect | 0.0: 706 (85.7%); 1.0: 73 (8.9%); 2.0: 21 (2.5%); 3.0: 13 (1.6%); 4.0: 11 (1.3%) | 0.0: 1159 (80.8%); 1.0: 149 (10.4%); 2.0: 70 (4.9%); 3.0: 21 (1.5%); 4.0: 36 (2.5%) | d = -0.133 | 0.002 | 0.003 |
| Abbreviations: HC, Healhty Controls; MDD, Major Depressive Disorder; ES, Effect Size; BMI, Body Mass Index; RSD-4, Recent Depressive Symptoms; CTS-5, Childhood Trauma Screener | | | | | |

**Table S4. Summary of stratification models' performances**

| **Model** | **N Clusters** | **Best SVM parameters** | **Stability (error)** | **Generalization accuracy** | **Sil (clusters)** | **DB (clusters)** | **p** |
| --- | --- | --- | --- | --- | --- | --- | --- |
| UKB training | 2 | C=0.1, kernel=linear | 0.12 (0.009) | / | 0.2 | 1.68 | <0.001 |
| UKB test | 2 | / | / | 0.965 | 0.2 | 1.71 | <0.001 |
| HSR validation | 2 | / | / | 0.806 | 0.32 | 1.38 | 0.008 |
| Abbreviations: UKB, UK Biobank; HSR, San Raffaele Hospital; SVM, support vector machine; Sil, Silhouette score; DB, Davies-Bouldin score | | | | | | | |

**Table S5. Comparisons on cortical thickness regional values (DKT atlas) between healthy controls and UKB clusters from the training set**

| **Variable** | **Groups (descriptive)** | **Test** | **Global p** | **FDR p** | **Effect size** | **Effect type** | **Post-hoc (FDR)** |
| --- | --- | --- | --- | --- | --- | --- | --- |
| Mean thickness of caudalanteriorcingulate (left hemisphere) | HC: 2.77 (2.57–2.94) \| CL1: 2.69 (2.53–2.84) \| CL2: 2.82 (2.63–2.99) | Kruskal–Wallis | <0.001 | <0.001 | 0.030 | Epsilon squared | HC vs CL1 (FDR p=<0.001); HC vs CL2 (FDR p=<0.001); CL1 vs CL2 (FDR p=<0.001) |
| Mean thickness of caudalanteriorcingulate (right hemisphere) | HC: 2.54 (2.33–2.75) \| CL1: 2.48 (2.25–2.63) \| CL2: 2.63 (2.42–2.83) | Kruskal–Wallis | <0.001 | <0.001 | 0.034 | Epsilon squared | HC vs CL1 (FDR p=<0.001); HC vs CL2 (FDR p=<0.001); CL1 vs CL2 (FDR p=<0.001) |
| Mean thickness of caudalmiddlefrontal (left hemisphere) | HC: 2.87 ± 0.15 \| CL1: 2.78 ± 0.14 \| CL2: 2.95 ± 0.11 | ANOVA | <0.001 | <0.001 | 0.159 | Eta squared | HC vs CL1 (FDR p=<0.001); HC vs CL2 (FDR p=<0.001); CL1 vs CL2 (FDR p=<0.001) |
| Mean thickness of caudalmiddlefrontal (right hemisphere) | HC: 2.83 ± 0.14 \| CL1: 2.74 ± 0.13 \| CL2: 2.90 ± 0.11 | ANOVA | <0.001 | <0.001 | 0.158 | Eta squared | HC vs CL1 (FDR p=<0.001); HC vs CL2 (FDR p=<0.001); CL1 vs CL2 (FDR p=<0.001) |
| Mean thickness of cuneus (left hemisphere) | HC: 2.04 ± 0.15 \| CL1: 1.97 ± 0.13 \| CL2: 2.10 ± 0.13 | ANOVA | <0.001 | <0.001 | 0.106 | Eta squared | HC vs CL1 (FDR p=<0.001); HC vs CL2 (FDR p=<0.001); CL1 vs CL2 (FDR p=<0.001) |
| Mean thickness of cuneus (right hemisphere) | HC: 1.95 ± 0.13 \| CL1: 1.88 ± 0.12 \| CL2: 2.00 ± 0.13 | ANOVA | <0.001 | <0.001 | 0.102 | Eta squared | HC vs CL1 (FDR p=<0.001); HC vs CL2 (FDR p=<0.001); CL1 vs CL2 (FDR p=<0.001) |
| Mean thickness of entorhinal (left hemisphere) | HC: 3.26 ± 0.28 \| CL1: 3.20 ± 0.28 \| CL2: 3.33 ± 0.26 | ANOVA | <0.001 | <0.001 | 0.027 | Eta squared | HC vs CL1 (FDR p=<0.001); HC vs CL2 (FDR p=<0.001); CL1 vs CL2 (FDR p=<0.001) |
| Mean thickness of entorhinal (right hemisphere) | HC: 3.36 ± 0.32 \| CL1: 3.26 ± 0.32 \| CL2: 3.41 ± 0.31 | ANOVA | <0.001 | <0.001 | 0.030 | Eta squared | HC vs CL1 (FDR p=<0.001); HC vs CL2 (FDR p=0.005); CL1 vs CL2 (FDR p=<0.001) |
| Mean thickness of fusiform (left hemisphere) | HC: 2.89 ± 0.14 \| CL1: 2.80 ± 0.12 \| CL2: 2.95 ± 0.10 | ANOVA | <0.001 | <0.001 | 0.163 | Eta squared | HC vs CL1 (FDR p=<0.001); HC vs CL2 (FDR p=<0.001); CL1 vs CL2 (FDR p=<0.001) |
| Mean thickness of fusiform (right hemisphere) | HC: 2.88 ± 0.14 \| CL1: 2.78 ± 0.13 \| CL2: 2.96 ± 0.12 | ANOVA | <0.001 | <0.001 | 0.176 | Eta squared | HC vs CL1 (FDR p=<0.001); HC vs CL2 (FDR p=<0.001); CL1 vs CL2 (FDR p=<0.001) |
| Mean thickness of inferiorparietal (left hemisphere) | HC: 2.71 (2.63–2.79) \| CL1: 2.64 (2.56–2.72) \| CL2: 2.77 (2.71–2.83) | Kruskal–Wallis | <0.001 | <0.001 | 0.161 | Epsilon squared | HC vs CL1 (FDR p=<0.001); HC vs CL2 (FDR p=<0.001); CL1 vs CL2 (FDR p=<0.001) |
| Mean thickness of inferiorparietal (right hemisphere) | HC: 2.74 ± 0.13 \| CL1: 2.65 ± 0.11 \| CL2: 2.81 ± 0.10 | ANOVA | <0.001 | <0.001 | 0.201 | Eta squared | HC vs CL1 (FDR p=<0.001); HC vs CL2 (FDR p=<0.001); CL1 vs CL2 (FDR p=<0.001) |
| Mean thickness of inferiortemporal (left hemisphere) | HC: 3.05 ± 0.14 \| CL1: 2.96 ± 0.14 \| CL2: 3.12 ± 0.12 | ANOVA | <0.001 | <0.001 | 0.136 | Eta squared | HC vs CL1 (FDR p=<0.001); HC vs CL2 (FDR p=<0.001); CL1 vs CL2 (FDR p=<0.001) |
| Mean thickness of inferiortemporal (right hemisphere) | HC: 3.03 ± 0.13 \| CL1: 2.94 ± 0.13 \| CL2: 3.08 ± 0.12 | ANOVA | <0.001 | <0.001 | 0.139 | Eta squared | HC vs CL1 (FDR p=<0.001); HC vs CL2 (FDR p=<0.001); CL1 vs CL2 (FDR p=<0.001) |
| Mean thickness of insula (left hemisphere) | HC: 3.21 ± 0.16 \| CL1: 3.11 ± 0.15 \| CL2: 3.26 ± 0.15 | ANOVA | <0.001 | <0.001 | 0.108 | Eta squared | HC vs CL1 (FDR p=<0.001); HC vs CL2 (FDR p=<0.001); CL1 vs CL2 (FDR p=<0.001) |
| Mean thickness of insula (right hemisphere) | HC: 3.20 ± 0.15 \| CL1: 3.10 ± 0.14 \| CL2: 3.26 ± 0.14 | ANOVA | <0.001 | <0.001 | 0.131 | Eta squared | HC vs CL1 (FDR p=<0.001); HC vs CL2 (FDR p=<0.001); CL1 vs CL2 (FDR p=<0.001) |
| Mean thickness of isthmuscingulate (left hemisphere) | HC: 2.48 ± 0.17 \| CL1: 2.43 ± 0.17 \| CL2: 2.53 ± 0.17 | ANOVA | <0.001 | <0.001 | 0.041 | Eta squared | HC vs CL1 (FDR p=<0.001); HC vs CL2 (FDR p=<0.001); CL1 vs CL2 (FDR p=<0.001) |
| Mean thickness of isthmuscingulate (right hemisphere) | HC: 2.54 ± 0.19 \| CL1: 2.47 ± 0.18 \| CL2: 2.61 ± 0.20 | ANOVA | <0.001 | <0.001 | 0.058 | Eta squared | HC vs CL1 (FDR p=<0.001); HC vs CL2 (FDR p=<0.001); CL1 vs CL2 (FDR p=<0.001) |
| Mean thickness of lateraloccipital (left hemisphere) | HC: 2.32 ± 0.13 \| CL1: 2.25 ± 0.11 \| CL2: 2.38 ± 0.11 | ANOVA | <0.001 | <0.001 | 0.125 | Eta squared | HC vs CL1 (FDR p=<0.001); HC vs CL2 (FDR p=<0.001); CL1 vs CL2 (FDR p=<0.001) |
| Mean thickness of lateraloccipital (right hemisphere) | HC: 2.36 ± 0.13 \| CL1: 2.27 ± 0.11 \| CL2: 2.42 ± 0.12 | ANOVA | <0.001 | <0.001 | 0.152 | Eta squared | HC vs CL1 (FDR p=<0.001); HC vs CL2 (FDR p=<0.001); CL1 vs CL2 (FDR p=<0.001) |
| Mean thickness of lateralorbitofrontal (left hemisphere) | HC: 2.83 ± 0.14 \| CL1: 2.74 ± 0.13 \| CL2: 2.90 ± 0.12 | ANOVA | <0.001 | <0.001 | 0.151 | Eta squared | HC vs CL1 (FDR p=<0.001); HC vs CL2 (FDR p=<0.001); CL1 vs CL2 (FDR p=<0.001) |
| Mean thickness of lateralorbitofrontal (right hemisphere) | HC: 2.82 ± 0.14 \| CL1: 2.73 ± 0.13 \| CL2: 2.89 ± 0.11 | ANOVA | <0.001 | <0.001 | 0.156 | Eta squared | HC vs CL1 (FDR p=<0.001); HC vs CL2 (FDR p=<0.001); CL1 vs CL2 (FDR p=<0.001) |
| Mean thickness of lingual (left hemisphere) | HC: 2.03 ± 0.15 \| CL1: 1.98 ± 0.13 \| CL2: 2.09 ± 0.14 | ANOVA | <0.001 | <0.001 | 0.077 | Eta squared | HC vs CL1 (FDR p=<0.001); HC vs CL2 (FDR p=<0.001); CL1 vs CL2 (FDR p=<0.001) |
| Mean thickness of lingual (right hemisphere) | HC: 2.01 ± 0.15 \| CL1: 1.95 ± 0.13 \| CL2: 2.06 ± 0.13 | ANOVA | <0.001 | <0.001 | 0.074 | Eta squared | HC vs CL1 (FDR p=<0.001); HC vs CL2 (FDR p=<0.001); CL1 vs CL2 (FDR p=<0.001) |
| Mean thickness of medialorbitofrontal (left hemisphere) | HC: 2.65 ± 0.17 \| CL1: 2.58 ± 0.17 \| CL2: 2.70 ± 0.16 | ANOVA | <0.001 | <0.001 | 0.069 | Eta squared | HC vs CL1 (FDR p=<0.001); HC vs CL2 (FDR p=<0.001); CL1 vs CL2 (FDR p=<0.001) |
| Mean thickness of medialorbitofrontal (right hemisphere) | HC: 2.70 (2.58–2.80) \| CL1: 2.58 (2.47–2.70) \| CL2: 2.75 (2.63–2.85) | Kruskal–Wallis | <0.001 | <0.001 | 0.099 | Epsilon squared | HC vs CL1 (FDR p=<0.001); HC vs CL2 (FDR p=<0.001); CL1 vs CL2 (FDR p=<0.001) |
| Mean thickness of middletemporal (left hemisphere) | HC: 2.90 ± 0.16 \| CL1: 2.82 ± 0.15 \| CL2: 2.97 ± 0.13 | ANOVA | <0.001 | <0.001 | 0.117 | Eta squared | HC vs CL1 (FDR p=<0.001); HC vs CL2 (FDR p=<0.001); CL1 vs CL2 (FDR p=<0.001) |
| Mean thickness of middletemporal (right hemisphere) | HC: 2.99 ± 0.15 \| CL1: 2.90 ± 0.13 \| CL2: 3.06 ± 0.13 | ANOVA | <0.001 | <0.001 | 0.150 | Eta squared | HC vs CL1 (FDR p=<0.001); HC vs CL2 (FDR p=<0.001); CL1 vs CL2 (FDR p=<0.001) |
| Mean thickness of paracentral (left hemisphere) | HC: 2.71 ± 0.19 \| CL1: 2.61 ± 0.18 \| CL2: 2.80 ± 0.15 | ANOVA | <0.001 | <0.001 | 0.134 | Eta squared | HC vs CL1 (FDR p=<0.001); HC vs CL2 (FDR p=<0.001); CL1 vs CL2 (FDR p=<0.001) |
| Mean thickness of paracentral (right hemisphere) | HC: 2.70 ± 0.19 \| CL1: 2.59 ± 0.16 \| CL2: 2.79 ± 0.15 | ANOVA | <0.001 | <0.001 | 0.145 | Eta squared | HC vs CL1 (FDR p=<0.001); HC vs CL2 (FDR p=<0.001); CL1 vs CL2 (FDR p=<0.001) |
| Mean thickness of parahippocampal (left hemisphere) | HC: 2.77 ± 0.30 \| CL1: 2.69 ± 0.31 \| CL2: 2.85 ± 0.30 | ANOVA | <0.001 | <0.001 | 0.031 | Eta squared | HC vs CL1 (FDR p=<0.001); HC vs CL2 (FDR p=<0.001); CL1 vs CL2 (FDR p=<0.001) |
| Mean thickness of parahippocampal (right hemisphere) | HC: 2.71 ± 0.26 \| CL1: 2.60 ± 0.25 \| CL2: 2.78 ± 0.24 | ANOVA | <0.001 | <0.001 | 0.060 | Eta squared | HC vs CL1 (FDR p=<0.001); HC vs CL2 (FDR p=<0.001); CL1 vs CL2 (FDR p=<0.001) |
| Mean thickness of parsopercularis (left hemisphere) | HC: 2.88 ± 0.15 \| CL1: 2.78 ± 0.15 \| CL2: 2.95 ± 0.12 | ANOVA | <0.001 | <0.001 | 0.136 | Eta squared | HC vs CL1 (FDR p=<0.001); HC vs CL2 (FDR p=<0.001); CL1 vs CL2 (FDR p=<0.001) |
| Mean thickness of parsopercularis (right hemisphere) | HC: 2.83 ± 0.14 \| CL1: 2.74 ± 0.14 \| CL2: 2.90 ± 0.12 | ANOVA | <0.001 | <0.001 | 0.150 | Eta squared | HC vs CL1 (FDR p=<0.001); HC vs CL2 (FDR p=<0.001); CL1 vs CL2 (FDR p=<0.001) |
| Mean thickness of parsorbitalis (left hemisphere) | HC: 2.88 ± 0.18 \| CL1: 2.78 ± 0.18 \| CL2: 2.96 ± 0.15 | ANOVA | <0.001 | <0.001 | 0.115 | Eta squared | HC vs CL1 (FDR p=<0.001); HC vs CL2 (FDR p=<0.001); CL1 vs CL2 (FDR p=<0.001) |
| Mean thickness of parsorbitalis (right hemisphere) | HC: 2.89 ± 0.19 \| CL1: 2.80 ± 0.17 \| CL2: 2.98 ± 0.16 | ANOVA | <0.001 | <0.001 | 0.118 | Eta squared | HC vs CL1 (FDR p=<0.001); HC vs CL2 (FDR p=<0.001); CL1 vs CL2 (FDR p=<0.001) |
| Mean thickness of parstriangularis (left hemisphere) | HC: 2.72 (2.62–2.81) \| CL1: 2.64 (2.55–2.72) \| CL2: 2.78 (2.70–2.85) | Kruskal–Wallis | <0.001 | <0.001 | 0.140 | Epsilon squared | HC vs CL1 (FDR p=<0.001); HC vs CL2 (FDR p=<0.001); CL1 vs CL2 (FDR p=<0.001) |
| Mean thickness of parstriangularis (right hemisphere) | HC: 2.69 ± 0.14 \| CL1: 2.59 ± 0.14 \| CL2: 2.75 ± 0.11 | ANOVA | <0.001 | <0.001 | 0.151 | Eta squared | HC vs CL1 (FDR p=<0.001); HC vs CL2 (FDR p=<0.001); CL1 vs CL2 (FDR p=<0.001) |
| Mean thickness of pericalcarine (left hemisphere) | HC: 1.73 ± 0.14 \| CL1: 1.68 ± 0.14 \| CL2: 1.79 ± 0.13 | ANOVA | <0.001 | <0.001 | 0.068 | Eta squared | HC vs CL1 (FDR p=<0.001); HC vs CL2 (FDR p=<0.001); CL1 vs CL2 (FDR p=<0.001) |
| Mean thickness of pericalcarine (right hemisphere) | HC: 1.70 ± 0.15 \| CL1: 1.63 ± 0.13 \| CL2: 1.75 ± 0.13 | ANOVA | <0.001 | <0.001 | 0.082 | Eta squared | HC vs CL1 (FDR p=<0.001); HC vs CL2 (FDR p=<0.001); CL1 vs CL2 (FDR p=<0.001) |
| Mean thickness of postcentral (left hemisphere) | HC: 2.35 ± 0.15 \| CL1: 2.27 ± 0.13 \| CL2: 2.43 ± 0.11 | ANOVA | <0.001 | <0.001 | 0.141 | Eta squared | HC vs CL1 (FDR p=<0.001); HC vs CL2 (FDR p=<0.001); CL1 vs CL2 (FDR p=<0.001) |
| Mean thickness of postcentral (right hemisphere) | HC: 2.31 ± 0.15 \| CL1: 2.22 ± 0.13 \| CL2: 2.38 ± 0.11 | ANOVA | <0.001 | <0.001 | 0.144 | Eta squared | HC vs CL1 (FDR p=<0.001); HC vs CL2 (FDR p=<0.001); CL1 vs CL2 (FDR p=<0.001) |
| Mean thickness of posteriorcingulate (left hemisphere) | HC: 2.69 ± 0.17 \| CL1: 2.63 ± 0.18 \| CL2: 2.74 ± 0.16 | ANOVA | <0.001 | <0.001 | 0.058 | Eta squared | HC vs CL1 (FDR p=<0.001); HC vs CL2 (FDR p=<0.001); CL1 vs CL2 (FDR p=<0.001) |
| Mean thickness of posteriorcingulate (right hemisphere) | HC: 2.70 ± 0.17 \| CL1: 2.62 ± 0.17 \| CL2: 2.76 ± 0.16 | ANOVA | <0.001 | <0.001 | 0.079 | Eta squared | HC vs CL1 (FDR p=<0.001); HC vs CL2 (FDR p=<0.001); CL1 vs CL2 (FDR p=<0.001) |
| Mean thickness of precentral (left hemisphere) | HC: 2.86 (2.74–2.96) \| CL1: 2.76 (2.63–2.85) \| CL2: 2.95 (2.85–3.03) | Kruskal–Wallis | <0.001 | <0.001 | 0.178 | Epsilon squared | HC vs CL1 (FDR p=<0.001); HC vs CL2 (FDR p=<0.001); CL1 vs CL2 (FDR p=<0.001) |
| Mean thickness of precentral (right hemisphere) | HC: 2.80 ± 0.17 \| CL1: 2.68 ± 0.15 \| CL2: 2.89 ± 0.14 | ANOVA | <0.001 | <0.001 | 0.176 | Eta squared | HC vs CL1 (FDR p=<0.001); HC vs CL2 (FDR p=<0.001); CL1 vs CL2 (FDR p=<0.001) |
| Mean thickness of precuneus (left hemisphere) | HC: 2.64 ± 0.14 \| CL1: 2.55 ± 0.13 \| CL2: 2.71 ± 0.10 | ANOVA | <0.001 | <0.001 | 0.172 | Eta squared | HC vs CL1 (FDR p=<0.001); HC vs CL2 (FDR p=<0.001); CL1 vs CL2 (FDR p=<0.001) |
| Mean thickness of precuneus (right hemisphere) | HC: 2.64 ± 0.13 \| CL1: 2.54 ± 0.11 \| CL2: 2.70 ± 0.10 | ANOVA | <0.001 | <0.001 | 0.193 | Eta squared | HC vs CL1 (FDR p=<0.001); HC vs CL2 (FDR p=<0.001); CL1 vs CL2 (FDR p=<0.001) |
| Mean thickness of rostralanteriorcingulate (left hemisphere) | HC: 2.90 ± 0.18 \| CL1: 2.82 ± 0.17 \| CL2: 2.96 ± 0.17 | ANOVA | <0.001 | <0.001 | 0.065 | Eta squared | HC vs CL1 (FDR p=<0.001); HC vs CL2 (FDR p=<0.001); CL1 vs CL2 (FDR p=<0.001) |
| Mean thickness of rostralanteriorcingulate (right hemisphere) | HC: 2.94 ± 0.20 \| CL1: 2.86 ± 0.18 \| CL2: 3.00 ± 0.20 | ANOVA | <0.001 | <0.001 | 0.061 | Eta squared | HC vs CL1 (FDR p=<0.001); HC vs CL2 (FDR p=<0.001); CL1 vs CL2 (FDR p=<0.001) |
| Mean thickness of rostralmiddlefrontal (left hemisphere) | HC: 2.70 (2.61–2.77) \| CL1: 2.62 (2.53–2.68) \| CL2: 2.75 (2.69–2.82) | Kruskal–Wallis | <0.001 | <0.001 | 0.175 | Epsilon squared | HC vs CL1 (FDR p=<0.001); HC vs CL2 (FDR p=<0.001); CL1 vs CL2 (FDR p=<0.001) |
| Mean thickness of rostralmiddlefrontal (right hemisphere) | HC: 2.65 (2.57–2.72) \| CL1: 2.57 (2.49–2.63) \| CL2: 2.70 (2.65–2.75) | Kruskal–Wallis | <0.001 | <0.001 | 0.202 | Epsilon squared | HC vs CL1 (FDR p=<0.001); HC vs CL2 (FDR p=<0.001); CL1 vs CL2 (FDR p=<0.001) |
| Mean thickness of superiorfrontal (left hemisphere) | HC: 2.96 ± 0.15 \| CL1: 2.86 ± 0.14 \| CL2: 3.04 ± 0.10 | ANOVA | <0.001 | <0.001 | 0.180 | Eta squared | HC vs CL1 (FDR p=<0.001); HC vs CL2 (FDR p=<0.001); CL1 vs CL2 (FDR p=<0.001) |
| Mean thickness of superiorfrontal (right hemisphere) | HC: 2.92 ± 0.13 \| CL1: 2.82 ± 0.13 \| CL2: 3.00 ± 0.09 | ANOVA | <0.001 | <0.001 | 0.213 | Eta squared | HC vs CL1 (FDR p=<0.001); HC vs CL2 (FDR p=<0.001); CL1 vs CL2 (FDR p=<0.001) |
| Mean thickness of superiorparietal (left hemisphere) | HC: 2.48 ± 0.14 \| CL1: 2.41 ± 0.13 \| CL2: 2.56 ± 0.10 | ANOVA | <0.001 | <0.001 | 0.160 | Eta squared | HC vs CL1 (FDR p=<0.001); HC vs CL2 (FDR p=<0.001); CL1 vs CL2 (FDR p=<0.001) |
| Mean thickness of superiorparietal (right hemisphere) | HC: 2.46 ± 0.13 \| CL1: 2.37 ± 0.12 \| CL2: 2.54 ± 0.10 | ANOVA | <0.001 | <0.001 | 0.187 | Eta squared | HC vs CL1 (FDR p=<0.001); HC vs CL2 (FDR p=<0.001); CL1 vs CL2 (FDR p=<0.001) |
| Mean thickness of superiortemporal (left hemisphere) | HC: 2.99 ± 0.17 \| CL1: 2.89 ± 0.15 \| CL2: 3.07 ± 0.14 | ANOVA | <0.001 | <0.001 | 0.136 | Eta squared | HC vs CL1 (FDR p=<0.001); HC vs CL2 (FDR p=<0.001); CL1 vs CL2 (FDR p=<0.001) |
| Mean thickness of superiortemporal (right hemisphere) | HC: 3.08 ± 0.16 \| CL1: 2.96 ± 0.14 \| CL2: 3.16 ± 0.13 | ANOVA | <0.001 | <0.001 | 0.178 | Eta squared | HC vs CL1 (FDR p=<0.001); HC vs CL2 (FDR p=<0.001); CL1 vs CL2 (FDR p=<0.001) |
| Mean thickness of supramarginal (left hemisphere) | HC: 2.84 (2.74–2.92) \| CL1: 2.76 (2.67–2.83) \| CL2: 2.91 (2.84–2.97) | Kruskal–Wallis | <0.001 | <0.001 | 0.181 | Epsilon squared | HC vs CL1 (FDR p=<0.001); HC vs CL2 (FDR p=<0.001); CL1 vs CL2 (FDR p=<0.001) |
| Mean thickness of supramarginal (right hemisphere) | HC: 2.85 (2.76–2.92) \| CL1: 2.74 (2.67–2.81) \| CL2: 2.91 (2.84–2.99) | Kruskal–Wallis | <0.001 | <0.001 | 0.212 | Epsilon squared | HC vs CL1 (FDR p=<0.001); HC vs CL2 (FDR p=<0.001); CL1 vs CL2 (FDR p=<0.001) |
| Mean thickness of transversetemporal (left hemisphere) | HC: 2.56 ± 0.25 \| CL1: 2.45 ± 0.23 \| CL2: 2.64 ± 0.24 | ANOVA | <0.001 | <0.001 | 0.078 | Eta squared | HC vs CL1 (FDR p=<0.001); HC vs CL2 (FDR p=<0.001); CL1 vs CL2 (FDR p=<0.001) |
| Mean thickness of transversetemporal (right hemisphere) | HC: 2.59 ± 0.26 \| CL1: 2.48 ± 0.24 \| CL2: 2.69 ± 0.25 | ANOVA | <0.001 | <0.001 | 0.078 | Eta squared | HC vs CL1 (FDR p=<0.001); HC vs CL2 (FDR p=<0.001); CL1 vs CL2 (FDR p=<0.001) |
| Abbreviations: HC, Healthy Controls; CL1, Cluster 1; CL2, Cluster 2 | | | | | | | |

**Table S6. Comparisons on cortical thickness regional values (DKT atlas) between healthy controls and UKB clusters from the test set**

| **Variable** | **Groups (descriptive)** | **Test** | **Global p** | **FDR p** | **Effect size** | **Effect type** | **Post-hoc (FDR)** |
| --- | --- | --- | --- | --- | --- | --- | --- |
| Mean thickness of caudalanteriorcingulate (left hemisphere) | HC: 2.77 (2.57–2.94) \| CL1: 2.67 (2.46–2.83) \| CL2: 2.82 (2.65–3.00) | Kruskal–Wallis | <0.001 | <0.001 | 0.039 | Epsilon squared | HC vs CL1 (FDR p=<0.001); HC vs CL2 (FDR p=<0.001); CL1 vs CL2 (FDR p=<0.001) |
| Mean thickness of caudalanteriorcingulate (right hemisphere) | HC: 2.53 ± 0.33 \| CL1: 2.48 ± 0.32 \| CL2: 2.60 ± 0.32 | ANOVA | <0.001 | <0.001 | 0.016 | Eta squared | HC vs CL1 (FDR p=0.025); HC vs CL2 (FDR p=<0.001); CL1 vs CL2 (FDR p=<0.001) |
| Mean thickness of caudalmiddlefrontal (left hemisphere) | HC: 2.87 ± 0.15 \| CL1: 2.77 ± 0.13 \| CL2: 2.94 ± 0.12 | ANOVA | <0.001 | <0.001 | 0.136 | Eta squared | HC vs CL1 (FDR p=<0.001); HC vs CL2 (FDR p=<0.001); CL1 vs CL2 (FDR p=<0.001) |
| Mean thickness of caudalmiddlefrontal (right hemisphere) | HC: 2.83 ± 0.14 \| CL1: 2.73 ± 0.14 \| CL2: 2.90 ± 0.11 | ANOVA | <0.001 | <0.001 | 0.143 | Eta squared | HC vs CL1 (FDR p=<0.001); HC vs CL2 (FDR p=<0.001); CL1 vs CL2 (FDR p=<0.001) |
| Mean thickness of cuneus (left hemisphere) | HC: 2.04 ± 0.15 \| CL1: 1.96 ± 0.13 \| CL2: 2.11 ± 0.13 | ANOVA | <0.001 | <0.001 | 0.104 | Eta squared | HC vs CL1 (FDR p=<0.001); HC vs CL2 (FDR p=<0.001); CL1 vs CL2 (FDR p=<0.001) |
| Mean thickness of cuneus (right hemisphere) | HC: 1.95 ± 0.13 \| CL1: 1.89 ± 0.12 \| CL2: 2.01 ± 0.13 | ANOVA | <0.001 | <0.001 | 0.078 | Eta squared | HC vs CL1 (FDR p=<0.001); HC vs CL2 (FDR p=<0.001); CL1 vs CL2 (FDR p=<0.001) |
| Mean thickness of entorhinal (left hemisphere) | HC: 3.26 ± 0.28 \| CL1: 3.12 ± 0.30 \| CL2: 3.32 ± 0.26 | ANOVA | <0.001 | <0.001 | 0.051 | Eta squared | HC vs CL1 (FDR p=<0.001); HC vs CL2 (FDR p=<0.001); CL1 vs CL2 (FDR p=<0.001) |
| Mean thickness of entorhinal (right hemisphere) | HC: 3.36 ± 0.32 \| CL1: 3.23 ± 0.33 \| CL2: 3.42 ± 0.31 | ANOVA | <0.001 | <0.001 | 0.037 | Eta squared | HC vs CL1 (FDR p=<0.001); HC vs CL2 (FDR p=0.003); CL1 vs CL2 (FDR p=<0.001) |
| Mean thickness of fusiform (left hemisphere) | HC: 2.89 ± 0.14 \| CL1: 2.80 ± 0.13 \| CL2: 2.95 ± 0.11 | ANOVA | <0.001 | <0.001 | 0.132 | Eta squared | HC vs CL1 (FDR p=<0.001); HC vs CL2 (FDR p=<0.001); CL1 vs CL2 (FDR p=<0.001) |
| Mean thickness of fusiform (right hemisphere) | HC: 2.88 ± 0.14 \| CL1: 2.79 ± 0.13 \| CL2: 2.95 ± 0.11 | ANOVA | <0.001 | <0.001 | 0.13 | Eta squared | HC vs CL1 (FDR p=<0.001); HC vs CL2 (FDR p=<0.001); CL1 vs CL2 (FDR p=<0.001) |
| Mean thickness of inferiorparietal (left hemisphere) | HC: 2.71 (2.63–2.79) \| CL1: 2.63 (2.58–2.69) \| CL2: 2.77 (2.71–2.83) | Kruskal–Wallis | <0.001 | <0.001 | 0.178 | Epsilon squared | HC vs CL1 (FDR p=<0.001); HC vs CL2 (FDR p=<0.001); CL1 vs CL2 (FDR p=<0.001) |
| Mean thickness of inferiorparietal (right hemisphere) | HC: 2.74 ± 0.13 \| CL1: 2.65 ± 0.11 \| CL2: 2.80 ± 0.10 | ANOVA | <0.001 | <0.001 | 0.161 | Eta squared | HC vs CL1 (FDR p=<0.001); HC vs CL2 (FDR p=<0.001); CL1 vs CL2 (FDR p=<0.001) |
| Mean thickness of inferiortemporal (left hemisphere) | HC: 3.05 ± 0.14 \| CL1: 2.96 ± 0.14 \| CL2: 3.11 ± 0.12 | ANOVA | <0.001 | <0.001 | 0.118 | Eta squared | HC vs CL1 (FDR p=<0.001); HC vs CL2 (FDR p=<0.001); CL1 vs CL2 (FDR p=<0.001) |
| Mean thickness of inferiortemporal (right hemisphere) | HC: 3.03 ± 0.13 \| CL1: 2.93 ± 0.12 \| CL2: 3.08 ± 0.12 | ANOVA | <0.001 | <0.001 | 0.133 | Eta squared | HC vs CL1 (FDR p=<0.001); HC vs CL2 (FDR p=<0.001); CL1 vs CL2 (FDR p=<0.001) |
| Mean thickness of insula (left hemisphere) | HC: 3.21 ± 0.16 \| CL1: 3.12 ± 0.15 \| CL2: 3.26 ± 0.14 | ANOVA | <0.001 | <0.001 | 0.09 | Eta squared | HC vs CL1 (FDR p=<0.001); HC vs CL2 (FDR p=<0.001); CL1 vs CL2 (FDR p=<0.001) |
| Mean thickness of insula (right hemisphere) | HC: 3.20 ± 0.15 \| CL1: 3.10 ± 0.15 \| CL2: 3.25 ± 0.14 | ANOVA | <0.001 | <0.001 | 0.107 | Eta squared | HC vs CL1 (FDR p=<0.001); HC vs CL2 (FDR p=<0.001); CL1 vs CL2 (FDR p=<0.001) |
| Mean thickness of isthmuscingulate (left hemisphere) | HC: 2.48 ± 0.17 \| CL1: 2.42 ± 0.18 \| CL2: 2.53 ± 0.16 | ANOVA | <0.001 | <0.001 | 0.036 | Eta squared | HC vs CL1 (FDR p=<0.001); HC vs CL2 (FDR p=<0.001); CL1 vs CL2 (FDR p=<0.001) |
| Mean thickness of isthmuscingulate (right hemisphere) | HC: 2.54 ± 0.19 \| CL1: 2.47 ± 0.17 \| CL2: 2.60 ± 0.19 | ANOVA | <0.001 | <0.001 | 0.049 | Eta squared | HC vs CL1 (FDR p=<0.001); HC vs CL2 (FDR p=<0.001); CL1 vs CL2 (FDR p=<0.001) |
| Mean thickness of lateraloccipital (left hemisphere) | HC: 2.32 ± 0.13 \| CL1: 2.25 ± 0.10 \| CL2: 2.37 ± 0.11 | ANOVA | <0.001 | <0.001 | 0.11 | Eta squared | HC vs CL1 (FDR p=<0.001); HC vs CL2 (FDR p=<0.001); CL1 vs CL2 (FDR p=<0.001) |
| Mean thickness of lateraloccipital (right hemisphere) | HC: 2.36 ± 0.13 \| CL1: 2.27 ± 0.12 \| CL2: 2.41 ± 0.12 | ANOVA | <0.001 | <0.001 | 0.12 | Eta squared | HC vs CL1 (FDR p=<0.001); HC vs CL2 (FDR p=<0.001); CL1 vs CL2 (FDR p=<0.001) |
| Mean thickness of lateralorbitofrontal (left hemisphere) | HC: 2.83 ± 0.14 \| CL1: 2.74 ± 0.14 \| CL2: 2.89 ± 0.12 | ANOVA | <0.001 | <0.001 | 0.107 | Eta squared | HC vs CL1 (FDR p=<0.001); HC vs CL2 (FDR p=<0.001); CL1 vs CL2 (FDR p=<0.001) |
| Mean thickness of lateralorbitofrontal (right hemisphere) | HC: 2.82 ± 0.14 \| CL1: 2.73 ± 0.14 \| CL2: 2.87 ± 0.13 | ANOVA | <0.001 | <0.001 | 0.106 | Eta squared | HC vs CL1 (FDR p=<0.001); HC vs CL2 (FDR p=<0.001); CL1 vs CL2 (FDR p=<0.001) |
| Mean thickness of lingual (left hemisphere) | HC: 2.03 ± 0.15 \| CL1: 1.95 ± 0.13 \| CL2: 2.08 ± 0.14 | ANOVA | <0.001 | <0.001 | 0.082 | Eta squared | HC vs CL1 (FDR p=<0.001); HC vs CL2 (FDR p=<0.001); CL1 vs CL2 (FDR p=<0.001) |
| Mean thickness of lingual (right hemisphere) | HC: 2.01 ± 0.15 \| CL1: 1.96 ± 0.13 \| CL2: 2.06 ± 0.14 | ANOVA | <0.001 | <0.001 | 0.052 | Eta squared | HC vs CL1 (FDR p=<0.001); HC vs CL2 (FDR p=<0.001); CL1 vs CL2 (FDR p=<0.001) |
| Mean thickness of medialorbitofrontal (left hemisphere) | HC: 2.65 ± 0.17 \| CL1: 2.56 ± 0.17 \| CL2: 2.69 ± 0.15 | ANOVA | <0.001 | <0.001 | 0.066 | Eta squared | HC vs CL1 (FDR p=<0.001); HC vs CL2 (FDR p=<0.001); CL1 vs CL2 (FDR p=<0.001) |
| Mean thickness of medialorbitofrontal (right hemisphere) | HC: 2.70 (2.58–2.80) \| CL1: 2.60 (2.49–2.69) \| CL2: 2.73 (2.63–2.84) | Kruskal–Wallis | <0.001 | <0.001 | 0.074 | Epsilon squared | HC vs CL1 (FDR p=<0.001); HC vs CL2 (FDR p=<0.001); CL1 vs CL2 (FDR p=<0.001) |
| Mean thickness of middletemporal (left hemisphere) | HC: 2.90 ± 0.16 \| CL1: 2.81 ± 0.14 \| CL2: 2.97 ± 0.13 | ANOVA | <0.001 | <0.001 | 0.119 | Eta squared | HC vs CL1 (FDR p=<0.001); HC vs CL2 (FDR p=<0.001); CL1 vs CL2 (FDR p=<0.001) |
| Mean thickness of middletemporal (right hemisphere) | HC: 2.99 ± 0.15 \| CL1: 2.89 ± 0.13 \| CL2: 3.05 ± 0.12 | ANOVA | <0.001 | <0.001 | 0.141 | Eta squared | HC vs CL1 (FDR p=<0.001); HC vs CL2 (FDR p=<0.001); CL1 vs CL2 (FDR p=<0.001) |
| Mean thickness of paracentral (left hemisphere) | HC: 2.71 ± 0.19 \| CL1: 2.59 ± 0.17 \| CL2: 2.80 ± 0.15 | ANOVA | <0.001 | <0.001 | 0.135 | Eta squared | HC vs CL1 (FDR p=<0.001); HC vs CL2 (FDR p=<0.001); CL1 vs CL2 (FDR p=<0.001) |
| Mean thickness of paracentral (right hemisphere) | HC: 2.70 ± 0.19 \| CL1: 2.59 ± 0.16 \| CL2: 2.79 ± 0.16 | ANOVA | <0.001 | <0.001 | 0.122 | Eta squared | HC vs CL1 (FDR p=<0.001); HC vs CL2 (FDR p=<0.001); CL1 vs CL2 (FDR p=<0.001) |
| Mean thickness of parahippocampal (left hemisphere) | HC: 2.77 ± 0.30 \| CL1: 2.70 ± 0.29 \| CL2: 2.85 ± 0.26 | ANOVA | <0.001 | <0.001 | 0.033 | Eta squared | HC vs CL1 (FDR p=<0.001); HC vs CL2 (FDR p=<0.001); CL1 vs CL2 (FDR p=<0.001) |
| Mean thickness of parahippocampal (right hemisphere) | HC: 2.71 ± 0.26 \| CL1: 2.63 ± 0.25 \| CL2: 2.78 ± 0.24 | ANOVA | <0.001 | <0.001 | 0.038 | Eta squared | HC vs CL1 (FDR p=<0.001); HC vs CL2 (FDR p=<0.001); CL1 vs CL2 (FDR p=<0.001) |
| Mean thickness of parsopercularis (left hemisphere) | HC: 2.88 ± 0.15 \| CL1: 2.78 ± 0.14 \| CL2: 2.93 ± 0.12 | ANOVA | <0.001 | <0.001 | 0.107 | Eta squared | HC vs CL1 (FDR p=<0.001); HC vs CL2 (FDR p=<0.001); CL1 vs CL2 (FDR p=<0.001) |
| Mean thickness of parsopercularis (right hemisphere) | HC: 2.83 ± 0.14 \| CL1: 2.73 ± 0.14 \| CL2: 2.88 ± 0.11 | ANOVA | <0.001 | <0.001 | 0.125 | Eta squared | HC vs CL1 (FDR p=<0.001); HC vs CL2 (FDR p=<0.001); CL1 vs CL2 (FDR p=<0.001) |
| Mean thickness of parsorbitalis (left hemisphere) | HC: 2.88 ± 0.18 \| CL1: 2.78 ± 0.16 \| CL2: 2.96 ± 0.16 | ANOVA | <0.001 | <0.001 | 0.102 | Eta squared | HC vs CL1 (FDR p=<0.001); HC vs CL2 (FDR p=<0.001); CL1 vs CL2 (FDR p=<0.001) |
| Mean thickness of parsorbitalis (right hemisphere) | HC: 2.89 ± 0.19 \| CL1: 2.78 ± 0.17 \| CL2: 2.97 ± 0.16 | ANOVA | <0.001 | <0.001 | 0.103 | Eta squared | HC vs CL1 (FDR p=<0.001); HC vs CL2 (FDR p=<0.001); CL1 vs CL2 (FDR p=<0.001) |
| Mean thickness of parstriangularis (left hemisphere) | HC: 2.71 ± 0.15 \| CL1: 2.61 ± 0.13 \| CL2: 2.77 ± 0.11 | ANOVA | <0.001 | <0.001 | 0.129 | Eta squared | HC vs CL1 (FDR p=<0.001); HC vs CL2 (FDR p=<0.001); CL1 vs CL2 (FDR p=<0.001) |
| Mean thickness of parstriangularis (right hemisphere) | HC: 2.69 ± 0.14 \| CL1: 2.59 ± 0.12 \| CL2: 2.75 ± 0.11 | ANOVA | <0.001 | <0.001 | 0.153 | Eta squared | HC vs CL1 (FDR p=<0.001); HC vs CL2 (FDR p=<0.001); CL1 vs CL2 (FDR p=<0.001) |
| Mean thickness of pericalcarine (left hemisphere) | HC: 1.73 ± 0.14 \| CL1: 1.67 ± 0.14 \| CL2: 1.79 ± 0.13 | ANOVA | <0.001 | <0.001 | 0.071 | Eta squared | HC vs CL1 (FDR p=<0.001); HC vs CL2 (FDR p=<0.001); CL1 vs CL2 (FDR p=<0.001) |
| Mean thickness of pericalcarine (right hemisphere) | HC: 1.70 ± 0.15 \| CL1: 1.65 ± 0.13 \| CL2: 1.76 ± 0.14 | ANOVA | <0.001 | <0.001 | 0.055 | Eta squared | HC vs CL1 (FDR p=<0.001); HC vs CL2 (FDR p=<0.001); CL1 vs CL2 (FDR p=<0.001) |
| Mean thickness of postcentral (left hemisphere) | HC: 2.35 ± 0.15 \| CL1: 2.27 ± 0.12 \| CL2: 2.43 ± 0.11 | ANOVA | <0.001 | <0.001 | 0.134 | Eta squared | HC vs CL1 (FDR p=<0.001); HC vs CL2 (FDR p=<0.001); CL1 vs CL2 (FDR p=<0.001) |
| Mean thickness of postcentral (right hemisphere) | HC: 2.31 ± 0.15 \| CL1: 2.23 ± 0.13 \| CL2: 2.38 ± 0.12 | ANOVA | <0.001 | <0.001 | 0.119 | Eta squared | HC vs CL1 (FDR p=<0.001); HC vs CL2 (FDR p=<0.001); CL1 vs CL2 (FDR p=<0.001) |
| Mean thickness of posteriorcingulate (left hemisphere) | HC: 2.69 ± 0.17 \| CL1: 2.62 ± 0.17 \| CL2: 2.74 ± 0.15 | ANOVA | <0.001 | <0.001 | 0.055 | Eta squared | HC vs CL1 (FDR p=<0.001); HC vs CL2 (FDR p=<0.001); CL1 vs CL2 (FDR p=<0.001) |
| Mean thickness of posteriorcingulate (right hemisphere) | HC: 2.70 ± 0.17 \| CL1: 2.64 ± 0.17 \| CL2: 2.75 ± 0.17 | ANOVA | <0.001 | <0.001 | 0.049 | Eta squared | HC vs CL1 (FDR p=<0.001); HC vs CL2 (FDR p=<0.001); CL1 vs CL2 (FDR p=<0.001) |
| Mean thickness of precentral (left hemisphere) | HC: 2.86 (2.74–2.96) \| CL1: 2.73 (2.63–2.82) \| CL2: 2.92 (2.85–3.00) | Kruskal–Wallis | <0.001 | <0.001 | 0.164 | Epsilon squared | HC vs CL1 (FDR p=<0.001); HC vs CL2 (FDR p=<0.001); CL1 vs CL2 (FDR p=<0.001) |
| Mean thickness of precentral (right hemisphere) | HC: 2.80 ± 0.17 \| CL1: 2.68 ± 0.16 \| CL2: 2.88 ± 0.14 | ANOVA | <0.001 | <0.001 | 0.15 | Eta squared | HC vs CL1 (FDR p=<0.001); HC vs CL2 (FDR p=<0.001); CL1 vs CL2 (FDR p=<0.001) |
| Mean thickness of precuneus (left hemisphere) | HC: 2.64 ± 0.14 \| CL1: 2.54 ± 0.13 \| CL2: 2.71 ± 0.10 | ANOVA | <0.001 | <0.001 | 0.167 | Eta squared | HC vs CL1 (FDR p=<0.001); HC vs CL2 (FDR p=<0.001); CL1 vs CL2 (FDR p=<0.001) |
| Mean thickness of precuneus (right hemisphere) | HC: 2.64 ± 0.13 \| CL1: 2.54 ± 0.11 \| CL2: 2.71 ± 0.10 | ANOVA | <0.001 | <0.001 | 0.165 | Eta squared | HC vs CL1 (FDR p=<0.001); HC vs CL2 (FDR p=<0.001); CL1 vs CL2 (FDR p=<0.001) |
| Mean thickness of rostralanteriorcingulate (left hemisphere) | HC: 2.90 ± 0.18 \| CL1: 2.82 ± 0.18 \| CL2: 2.95 ± 0.17 | ANOVA | <0.001 | <0.001 | 0.056 | Eta squared | HC vs CL1 (FDR p=<0.001); HC vs CL2 (FDR p=<0.001); CL1 vs CL2 (FDR p=<0.001) |
| Mean thickness of rostralanteriorcingulate (right hemisphere) | HC: 2.94 ± 0.20 \| CL1: 2.86 ± 0.20 \| CL2: 2.99 ± 0.17 | ANOVA | <0.001 | <0.001 | 0.044 | Eta squared | HC vs CL1 (FDR p=<0.001); HC vs CL2 (FDR p=<0.001); CL1 vs CL2 (FDR p=<0.001) |
| Mean thickness of rostralmiddlefrontal (left hemisphere) | HC: 2.69 ± 0.13 \| CL1: 2.61 ± 0.11 \| CL2: 2.75 ± 0.10 | ANOVA | <0.001 | <0.001 | 0.138 | Eta squared | HC vs CL1 (FDR p=<0.001); HC vs CL2 (FDR p=<0.001); CL1 vs CL2 (FDR p=<0.001) |
| Mean thickness of rostralmiddlefrontal (right hemisphere) | HC: 2.65 (2.57–2.72) \| CL1: 2.58 (2.51–2.63) \| CL2: 2.68 (2.64–2.75) | Kruskal–Wallis | <0.001 | <0.001 | 0.141 | Epsilon squared | HC vs CL1 (FDR p=<0.001); HC vs CL2 (FDR p=<0.001); CL1 vs CL2 (FDR p=<0.001) |
| Mean thickness of superiorfrontal (left hemisphere) | HC: 2.96 ± 0.15 \| CL1: 2.86 ± 0.13 \| CL2: 3.03 ± 0.10 | ANOVA | <0.001 | <0.001 | 0.162 | Eta squared | HC vs CL1 (FDR p=<0.001); HC vs CL2 (FDR p=<0.001); CL1 vs CL2 (FDR p=<0.001) |
| Mean thickness of superiorfrontal (right hemisphere) | HC: 2.93 (2.84–3.01) \| CL1: 2.84 (2.77–2.91) \| CL2: 2.99 (2.93–3.05) | Kruskal–Wallis | <0.001 | <0.001 | 0.172 | Epsilon squared | HC vs CL1 (FDR p=<0.001); HC vs CL2 (FDR p=<0.001); CL1 vs CL2 (FDR p=<0.001) |
| Mean thickness of superiorparietal (left hemisphere) | HC: 2.50 (2.40–2.58) \| CL1: 2.42 (2.33–2.48) \| CL2: 2.56 (2.49–2.62) | Kruskal–Wallis | <0.001 | <0.001 | 0.17 | Epsilon squared | HC vs CL1 (FDR p=<0.001); HC vs CL2 (FDR p=<0.001); CL1 vs CL2 (FDR p=<0.001) |
| Mean thickness of superiorparietal (right hemisphere) | HC: 2.46 ± 0.13 \| CL1: 2.37 ± 0.12 \| CL2: 2.53 ± 0.11 | ANOVA | <0.001 | <0.001 | 0.151 | Eta squared | HC vs CL1 (FDR p=<0.001); HC vs CL2 (FDR p=<0.001); CL1 vs CL2 (FDR p=<0.001) |
| Mean thickness of superiortemporal (left hemisphere) | HC: 2.99 ± 0.17 \| CL1: 2.89 ± 0.13 \| CL2: 3.06 ± 0.13 | ANOVA | <0.001 | <0.001 | 0.124 | Eta squared | HC vs CL1 (FDR p=<0.001); HC vs CL2 (FDR p=<0.001); CL1 vs CL2 (FDR p=<0.001) |
| Mean thickness of superiortemporal (right hemisphere) | HC: 3.08 ± 0.16 \| CL1: 2.96 ± 0.14 \| CL2: 3.14 ± 0.12 | ANOVA | <0.001 | <0.001 | 0.148 | Eta squared | HC vs CL1 (FDR p=<0.001); HC vs CL2 (FDR p=<0.001); CL1 vs CL2 (FDR p=<0.001) |
| Mean thickness of supramarginal (left hemisphere) | HC: 2.84 (2.74–2.92) \| CL1: 2.73 (2.66–2.81) \| CL2: 2.90 (2.84–2.97) | Kruskal–Wallis | <0.001 | <0.001 | 0.186 | Epsilon squared | HC vs CL1 (FDR p=<0.001); HC vs CL2 (FDR p=<0.001); CL1 vs CL2 (FDR p=<0.001) |
| Mean thickness of supramarginal (right hemisphere) | HC: 2.85 (2.76–2.92) \| CL1: 2.74 (2.66–2.82) \| CL2: 2.90 (2.83–2.97) | Kruskal–Wallis | <0.001 | <0.001 | 0.162 | Epsilon squared | HC vs CL1 (FDR p=<0.001); HC vs CL2 (FDR p=<0.001); CL1 vs CL2 (FDR p=<0.001) |
| Mean thickness of transversetemporal (left hemisphere) | HC: 2.56 ± 0.25 \| CL1: 2.44 ± 0.23 \| CL2: 2.63 ± 0.23 | ANOVA | <0.001 | <0.001 | 0.064 | Eta squared | HC vs CL1 (FDR p=<0.001); HC vs CL2 (FDR p=<0.001); CL1 vs CL2 (FDR p=<0.001) |
| Mean thickness of transversetemporal (right hemisphere) | HC: 2.59 ± 0.26 \| CL1: 2.48 ± 0.25 \| CL2: 2.68 ± 0.24 | ANOVA | <0.001 | <0.001 | 0.062 | Eta squared | HC vs CL1 (FDR p=<0.001); HC vs CL2 (FDR p=<0.001); CL1 vs CL2 (FDR p=<0.001) |
| Abbreviations: HC, Healthy Controls; CL1, Cluster 1; CL2, Cluster 2 | | | | | | | |

**Table S7. Comparisons on outcome variables between healthy controls and UKB clusters from the training set**

| **Variable** | **HC (N=827)** | **Cluster 1 (N=386)** | **Cluster 2 (N=456)** | **ES** | **p** | **q** | **Post-hoc (FDR)** |
| --- | --- | --- | --- | --- | --- | --- | --- |
| Age (years) | 58.00 (55.00–61.00) | 57.00 (53.00–61.00) | 58.00 (54.00–62.00) | η2 = 0.001 | 0.12 | 0.185 |  |
| Sex (% male) | 262/827 (31.7%) | 134/386 (34.7%) | 133/456 (29.2%) | V = 0.042 | 0.226 | 0.292 |  |
| BMI | 25.63 (23.27–28.43) | 27.20 (23.95–30.70) | 27.00 (23.70–30.58) | η2 = 0.016 | <0.001 | <0.001 | HC < CL1(FDR p=<0.001); HC < CL2 (FDR p=<0.001) |
| Education | 6.00 (4.00–6.00) | 5.00 (3.00–6.00) | 6.00 (4.00–6.00) | η2 = 0.001 | 0.126 | 0.185 |  |
| Age of onset (years) | / | 41.00 (31.00–49.00) | 41.00 (33.00–50.00) | d = -0.063 | 0.533 | 0.586 |  |
| Number of depressive episodes | / | 4.00 (2.00–9.00) | 4.00 (2.00–7.75) | d = -0.036 | 0.751 | 0.787 |  |
| Antidepressant treatment | / | 123/284 (43.3%) | 138/323 (42.7%) | V = 0.003 | 0.95 | 0.95 |  |
| Diabetes / cardiometabolic comorbidity | 32/827 (3.9%) | 42/386 (10.9%) | 29/455 (6.4%) | V = 0.116 | <0.001 | <0.001 | HC < CL1 (FDR p=<0.001); HC < CL2 (FDR p=<0.001); CL1 > CL2 (FDR p=0.018) |
| Depression with anxious features | / | 141/373 (37.8%) | 134/438 (30.6%) | V = 0.073 | 0.037 | 0.063 |  |
| Depression with atypical features | / | 45/367 (12.3%) | 41/432 (9.5%) | V = 0.041 | 0.252 | 0.294 |  |
| Treatment resistant depression | / | 20/153 (13.1%) | 14/165 (8.5%) | V = 0.064 | 0.254 | 0.294 |  |
| RDS-4 Total score | 1.00 (0.00–1.00) | 4.00 (3.00–6.00) | 4.00 (3.00–5.00) | η2 = 0.622 | <0.001 | <0.001 | HC < CL1(FDR p=<0.001); HC < CL2 (FDR p=<0.001); CL1 > CL2 (FDR p=0.029) |
| RSD-4 Depressed mood | 0: 827 (100.0%) | 0: 7 (1.8%); 1: 287 (74.4%); 2: 49 (12.7%); 3: 43 (11.1%) | 0: 9 (2.0%); 1: 343 (75.2%); 2: 53 (11.6%); 3: 51 (11.2%) | η2 = 0.883 | <0.001 | <0.001 | HC < CL1(FDR p=<0.001); HC < CL2 (FDR p=<0.001) |
| RSD-4 Lethargy | 0.0: 445 (54.7%); 1.0: 311 (38.3%); 2.0: 31 (3.8%); 3.0: 26 (3.2%) | 0.0: 115 (30.3%); 1.0: 200 (52.8%); 2.0: 33 (8.7%); 3.0: 31 (8.2%) | 0.0: 185 (41.4%); 1.0: 194 (43.4%); 2.0: 36 (8.1%); 3.0: 32 (7.2%) | η2 = 0.045 | <0.001 | <0.001 | HC < CL1(FDR p=<0.001); HC < CL2 (FDR p=<0.001); CL1 > CL2 (FDR p=0.005) |
| RSD-4 Tensensess | 0.0: 629 (77.7%); 1.0: 164 (20.2%); 2.0: 11 (1.4%); 3.0: 6 (0.7%) | 0.0: 47 (12.3%); 1.0: 199 (52.0%); 2.0: 53 (13.8%); 3.0: 84 (21.9%) | 0.0: 54 (12.0%); 1.0: 232 (51.4%); 2.0: 72 (16.0%); 3.0: 93 (20.6%) | η2 = 0.449 | <0.001 | <0.001 | HC < CL1(FDR p=<0.001); HC < CL2 (FDR p=<0.001) |
| RSD-4 Unenthusiasm | 0.0: 768 (93.7%); 1.0: 46 (5.6%); 2.0: 2 (0.2%); 3.0: 4 (0.5%) | 0.0: 84 (22.2%); 1.0: 222 (58.7%); 2.0: 40 (10.6%); 3.0: 32 (8.5%) | 0.0: 132 (29.3%); 1.0: 246 (54.5%); 2.0: 47 (10.4%); 3.0: 26 (5.8%) | η2 = 0.462 | <0.001 | <0.001 | HC < CL1(FDR p=<0.001); HC < CL2 (FDR p=<0.001); CL1 > CL2 (FDR p=0.023) |
| CTS-5 Total score | 1.00 (0.00–3.00) | 3.00 (1.00–5.00) | 2.00 (0.00–4.00) | η2 = 0.043 | <0.001 | <0.001 | HC < CL1(FDR p=<0.001); HC < CL2 (FDR p=<0.001) |
| CTS-5 Emotional Neglect | 0.0: 423 (51.2%); 1.0: 197 (23.8%); 2.0: 152 (18.4%); 3.0: 41 (5.0%); 4.0: 13 (1.6%) | 0.0: 129 (35.2%); 1.0: 80 (21.9%); 2.0: 102 (27.9%); 3.0: 39 (10.7%); 4.0: 16 (4.4%) | 0.0: 142 (32.9%); 1.0: 103 (23.9%); 2.0: 120 (27.8%); 3.0: 47 (10.9%); 4.0: 19 (4.4%) | η2 = 0.042 | <0.001 | <0.001 | HC < CL1(FDR p=<0.001); HC < CL2 (FDR p=<0.001) |
| CTS-5 Emotional Abuse | 0.0: 677 (82.1%); 1.0: 63 (7.6%); 2.0: 56 (6.8%); 3.0: 17 (2.1%); 4.0: 12 (1.5%) | 0.0: 224 (61.7%); 1.0: 38 (10.5%); 2.0: 62 (17.1%); 3.0: 22 (6.1%); 4.0: 17 (4.7%) | 0.0: 300 (69.8%); 1.0: 37 (8.6%); 2.0: 59 (13.7%); 3.0: 15 (3.5%); 4.0: 19 (4.4%) | η2 = 0.040 | <0.001 | <0.001 | HC < CL1 (FDR p=<0.001); HC < CL2 (FDR p=<0.001); CL1 > CL2 (FDR p=0.018) |
| CTS-5 Physical Abuse | 0.0: 617 (74.7%); 1.0: 126 (15.3%); 2.0: 65 (7.9%); 3.0: 8 (1.0%); 4.0: 10 (1.2%) | 0.0: 246 (67.4%); 1.0: 56 (15.3%); 2.0: 48 (13.2%); 3.0: 10 (2.7%); 4.0: 5 (1.4%) | 0.0: 309 (72.0%); 1.0: 55 (12.8%); 2.0: 49 (11.4%); 3.0: 9 (2.1%); 4.0: 7 (1.6%) | η2 = 0.004 | 0.012 | 0.022 | HC < CL1 (FDR p=0.009) |
| CTS-5 Sexual Abuse | 0.0: 734 (89.6%); 1.0: 35 (4.3%); 2.0: 35 (4.3%); 3.0: 10 (1.2%); 4.0: 5 (0.6%) | 0.0: 295 (81.5%); 1.0: 31 (8.6%); 2.0: 23 (6.4%); 3.0: 5 (1.4%); 4.0: 8 (2.2%) | 0.0: 366 (86.3%); 1.0: 23 (5.4%); 2.0: 29 (6.8%); 3.0: 4 (0.9%); 4.0: 2 (0.5%) | η2 = 0.008 | <0.001 | <0.001 | HC < CL1 (FDR p=<0.001) |
| CTS-5 Physical Neglect | 0.0: 706 (85.7%); 1.0: 73 (8.9%); 2.0: 21 (2.5%); 3.0: 13 (1.6%); 4.0: 11 (1.3%) | 0.0: 302 (82.7%); 1.0: 39 (10.7%); 2.0: 13 (3.6%); 3.0: 3 (0.8%); 4.0: 8 (2.2%) | 0.0: 351 (82.0%); 1.0: 43 (10.0%); 2.0: 17 (4.0%); 3.0: 8 (1.9%); 4.0: 9 (2.1%) | η2 = 0.001 | 0.165 | 0.227 |  |
| Abbreviations: HC, Healthy Controls; ES, Effect Size; BMI, Body Mass Index; RSD-4, Recent Depressive Symptoms; CTS-5, Childhood Trauma Screener | | | | | | | |

**Table S8. Comparisons on outcome variables between healthy controls and UKB clusters from the test set**

| **Variable** | **HC (N=827)** | **Cluster 1 (N=279)** | **Cluster 2 (N=410)** | **ES** | **p** | **q** | **Post-hoc (FDR)** |
| --- | --- | --- | --- | --- | --- | --- | --- |
| Age (years) | 58.00 (55.00–61.00) | 57.00 (53.00–61.00) | 58.00 (54.00–62.00) | η2 = 0.002 | 0.072 | 0.106 |  |
| Sex (% male) | 262/827 (31.7%) | 89/279 (31.9%) | 129/410 (31.5%) | V = 0.003 | 0.993 | 1 |  |
| BMI | 25.63 (23.27–28.43) | 27.00 (24.00–31.35) | 27.20 (23.90–30.90) | η2 = 0.021 | <0.001 | <0.001 | HC < CL1 (FDR p=<0.001); HC < CL2 (FDR p=<0.001) |
| Education | 6.00 (4.00–6.00) | 5.00 (3.00–6.00) | 5.00 (4.00–6.00) | η2 = 0.001 | 0.168 | 0.217 |  |
| Age of onset (years) | / | 40.00 (29.00–47.00) | 40.00 (30.00–49.00) | d = -0.040 | 0.694 | 0.763 |  |
| Number of depressive episodes | / | 4.00 (2.00–8.00) | 4.00 (2.00–6.00) | d = 0.070 | 0.129 | 0.177 |  |
| Antidepressant treatment | / | 92/208 (44.2%) | 114/293 (38.9%) | V = 0.049 | 0.271 | 0.331 |  |
| Diabetes / cardiometabolic comorbidity | 32/827 (3.9%) | 23/278 (8.3%) | 12/409 (2.9%) | V = 0.091 | 0.002 | 0.003 | HC < CL1 (FDR p<0.001); CL1 > CL2 (FDR p<0.001) |
| Depression with anxious features | / | 79/266 (29.7%) | 143/386 (37.0%) | V = 0.073 | 0.063 | 0.099 |  |
| Depression with atypical features | / | 27/265 (10.2%) | 38/381 (10.0%) | V = 0.000 | 1 | 1 |  |
| Treatment resistant depression | / | 12/112 (10.7%) | 18/135 (13.3%) | V = 0.027 | 0.666 | 0.763 |  |
| RDS-4 Total score | 1.00 (0.00–1.00) | 4.00 (3.00–6.00) | 4.00 (3.00–6.00) | η2 = 0.612 | <0.001 | <0.001 | HC < CL1 (FDR p=<0.001); HC < CL2 (FDR p=<0.001) |
| RSD-4 Depressed mood | 0: 827 (100.0%) | 0: 3 (1.1%); 1: 215 (77.1%); 2: 32 (11.5%); 3: 29 (10.4%) | 0: 10 (2.4%); 1: 306 (74.6%); 2: 41 (10.0%); 3: 53 (12.9%) | η2 = 0.907 | <0.001 | <0.001 | HC < CL1 (FDR p=<0.001); HC < CL2 (FDR p=<0.001) |
| RSD-4 Lethargy | 0.0: 445 (54.7%); 1.0: 311 (38.3%); 2.0: 31 (3.8%); 3.0: 26 (3.2%) | 0.0: 99 (35.9%); 1.0: 127 (46.0%); 2.0: 28 (10.1%); 3.0: 22 (8.0%) | 0.0: 126 (31.5%); 1.0: 206 (51.5%); 2.0: 28 (7.0%); 3.0: 40 (10.0%) | η2 = 0.054 | <0.001 | <0.001 | HC < CL1 (FDR p=<0.001); HC < CL2 (FDR p=<0.001) |
| RSD-4 Tensensess | 0.0: 629 (77.7%); 1.0: 164 (20.2%); 2.0: 11 (1.4%); 3.0: 6 (0.7%) | 0.0: 35 (12.6%); 1.0: 125 (45.0%); 2.0: 53 (19.1%); 3.0: 65 (23.4%) | 0.0: 57 (13.9%); 1.0: 193 (47.2%); 2.0: 68 (16.6%); 3.0: 91 (22.2%) | η2 = 0.448 | <0.001 | <0.001 | HC < CL1 (FDR p=<0.001); HC < CL2 (FDR p=<0.001) |
| RSD-4 Unenthusiasm | 0.0: 768 (93.7%); 1.0: 46 (5.6%); 2.0: 2 (0.2%); 3.0: 4 (0.5%) | 0.0: 66 (23.9%); 1.0: 156 (56.5%); 2.0: 32 (11.6%); 3.0: 22 (8.0%) | 0.0: 112 (28.0%); 1.0: 211 (52.8%); 2.0: 37 (9.2%); 3.0: 40 (10.0%) | η2 = 0.473 | <0.001 | <0.001 | HC < CL1 (FDR p=<0.001); HC < CL2 (FDR p=<0.001) |
| CTS-5 Total score | 1.00 (0.00–3.00) | 3.00 (1.00–5.25) | 2.00 (0.00–4.00) | η2 = 0.047 | <0.001 | <0.001 | HC < CL1 (FDR p=<0.001); HC < CL2 (FDR p=<0.001); CL1 > CL2 (FDR p=0.007) |
| CTS-5 Emotional Neglect | 0.0: 423 (51.2%); 1.0: 197 (23.8%); 2.0: 152 (18.4%); 3.0: 41 (5.0%); 4.0: 13 (1.6%) | 0.0: 70 (26.6%); 1.0: 63 (24.0%); 2.0: 81 (30.8%); 3.0: 34 (12.9%); 4.0: 15 (5.7%) | 0.0: 139 (36.8%); 1.0: 80 (21.2%); 2.0: 107 (28.3%); 3.0: 41 (10.8%); 4.0: 11 (2.9%) | η2 = 0.054 | <0.001 | <0.001 | HC < CL1 (FDR p=<0.001); HC < CL2 (FDR p=<0.001); CL1 > CL2 (FDR p=0.008) |
| CTS-5 Emotional Abuse | 0.0: 677 (82.1%); 1.0: 63 (7.6%); 2.0: 56 (6.8%); 3.0: 17 (2.1%); 4.0: 12 (1.5%) | 0.0: 163 (61.7%); 1.0: 33 (12.5%); 2.0: 33 (12.5%); 3.0: 20 (7.6%); 4.0: 15 (5.7%) | 0.0: 249 (65.7%); 1.0: 35 (9.2%); 2.0: 59 (15.6%); 3.0: 15 (4.0%); 4.0: 21 (5.5%) | η2 = 0.045 | <0.001 | <0.001 | HC < CL1 (FDR p=<0.001); HC < CL2 (FDR p=<0.001) |
| CTS-5 Physical Abuse | 0.0: 617 (74.7%); 1.0: 126 (15.3%); 2.0: 65 (7.9%); 3.0: 8 (1.0%); 4.0: 10 (1.2%) | 0.0: 165 (62.5%); 1.0: 42 (15.9%); 2.0: 45 (17.0%); 3.0: 7 (2.7%); 4.0: 5 (1.9%) | 0.0: 262 (68.8%); 1.0: 53 (13.9%); 2.0: 52 (13.6%); 3.0: 9 (2.4%); 4.0: 5 (1.3%) | η2 = 0.012 | <0.001 | <0.001 | HC < CL1 (FDR p=<0.001); HC < CL2 (FDR p=0.016) |
| CTS-5 Sexual Abuse | 0.0: 734 (89.6%); 1.0: 35 (4.3%); 2.0: 35 (4.3%); 3.0: 10 (1.2%); 4.0: 5 (0.6%) | 0.0: 213 (81.3%); 1.0: 19 (7.3%); 2.0: 20 (7.6%); 3.0: 3 (1.1%); 4.0: 7 (2.7%) | 0.0: 323 (88.3%); 1.0: 15 (4.1%); 2.0: 18 (4.9%); 3.0: 6 (1.6%); 4.0: 4 (1.1%) | η2 = 0.008 | 0.001 | 0.002 | HC < CL1 (FDR p=<0.001); CL1 < CL2 (FDR p=0.025) |
| CTS-5 Physical Neglect | 0.0: 706 (85.7%); 1.0: 73 (8.9%); 2.0: 21 (2.5%); 3.0: 13 (1.6%); 4.0: 11 (1.3%) | 0.0: 201 (76.4%); 1.0: 30 (11.4%); 2.0: 15 (5.7%); 3.0: 6 (2.3%); 4.0: 11 (4.2%) | 0.0: 305 (80.5%); 1.0: 37 (9.8%); 2.0: 25 (6.6%); 3.0: 4 (1.1%); 4.0: 8 (2.1%) | η2 = 0.009 | <0.001 | <0.001 | HC < CL1 (FDR p=<0.001); HC < CL2 (FDR p=0.026) |
| Abbreviations: HC, Healthy Controls; ES, Effect Size; BMI, Body Mass Index; RSD-4, Recent Depressive Symptoms; CTS-5, Childhood Trauma Screener | | | | | | | |

**Table S9. Weighted comparisons on cortical thickness regional values (DKT atlas) between clusters from HSR validation cohort**

| **Variable** | **Test** | **n (Cluster 1) [ESS]** | **n (Cluster 2) [ESS]** | **Cluster 1 (Mean ± SD)** | **Cluster 2 (Mean ± SD)** | **Effect Size** | **Effect type** | **Global p** | **FDR p** |
| --- | --- | --- | --- | --- | --- | --- | --- | --- | --- |
| Mean thickness of caudalanteriorcingulate (left hemisphere) | Weighted t-test (Cluster-robust SE) | 116 (ESS: 47.5) | 28 (ESS: 10.2) | 2.27 ± 0.15 | 2.50 ± 0.13 | -1.539 | Cohen's d | <0.001 | <0.001 |
| Mean thickness of caudalanteriorcingulate (right hemisphere) | Weighted t-test (Cluster-robust SE) | 116 (ESS: 47.5) | 28 (ESS: 10.2) | 2.11 ± 0.13 | 2.30 ± 0.16 | -1.384 | Cohen's d | <0.001 | <0.001 |
| Mean thickness of caudalmiddlefrontal (left hemisphere) | Weighted t-test (Cluster-robust SE) | 116 (ESS: 47.5) | 28 (ESS: 10.2) | 2.36 ± 0.10 | 2.49 ± 0.13 | -1.188 | Cohen's d | 0.001 | 0.002 |
| Mean thickness of caudalmiddlefrontal (right hemisphere) | Weighted t-test (Cluster-robust SE) | 116 (ESS: 47.5) | 28 (ESS: 10.2) | 2.31 ± 0.10 | 2.48 ± 0.09 | -1.778 | Cohen's d | <0.001 | <0.001 |
| Mean thickness of cuneus (left hemisphere) | Weighted t-test (Cluster-robust SE) | 116 (ESS: 47.5) | 28 (ESS: 10.2) | 1.85 ± 0.09 | 2.00 ± 0.11 | -1.5 | Cohen's d | <0.001 | <0.001 |
| Mean thickness of cuneus (right hemisphere) | Weighted t-test (Cluster-robust SE) | 116 (ESS: 47.5) | 28 (ESS: 10.2) | 1.85 ± 0.10 | 1.96 ± 0.12 | -1.09 | Cohen's d | 0.003 | 0.004 |
| Mean thickness of entorhinal (left hemisphere) | Weighted t-test (Cluster-robust SE) | 116 (ESS: 47.5) | 28 (ESS: 10.2) | 2.90 ± 0.22 | 3.28 ± 0.31 | -1.605 | Cohen's d | <0.001 | <0.001 |
| Mean thickness of entorhinal (right hemisphere) | Weighted t-test (Cluster-robust SE) | 116 (ESS: 47.5) | 28 (ESS: 10.2) | 3.02 ± 0.26 | 3.30 ± 0.41 | -0.992 | Cohen's d | 0.055 | 0.058 |
| Mean thickness of fusiform (left hemisphere) | Weighted t-test (Cluster-robust SE) | 116 (ESS: 47.5) | 28 (ESS: 10.2) | 2.45 ± 0.07 | 2.63 ± 0.08 | -2.495 | Cohen's d | <0.001 | <0.001 |
| Mean thickness of fusiform (right hemisphere) | Weighted t-test (Cluster-robust SE) | 116 (ESS: 47.5) | 28 (ESS: 10.2) | 2.46 ± 0.10 | 2.68 ± 0.07 | -2.354 | Cohen's d | <0.001 | <0.001 |
| Mean thickness of inferiorparietal (left hemisphere) | Weighted t-test (Cluster-robust SE) | 116 (ESS: 47.5) | 28 (ESS: 10.2) | 2.24 ± 0.07 | 2.47 ± 0.07 | -3.209 | Cohen's d | <0.001 | <0.001 |
| Mean thickness of inferiorparietal (right hemisphere) | Weighted t-test (Cluster-robust SE) | 116 (ESS: 47.5) | 28 (ESS: 10.2) | 2.24 ± 0.08 | 2.42 ± 0.08 | -2.381 | Cohen's d | <0.001 | <0.001 |
| Mean thickness of inferiortemporal (left hemisphere) | Weighted t-test (Cluster-robust SE) | 116 (ESS: 47.5) | 28 (ESS: 10.2) | 2.54 ± 0.09 | 2.73 ± 0.12 | -2.058 | Cohen's d | <0.001 | <0.001 |
| Mean thickness of inferiortemporal (right hemisphere) | Weighted t-test (Cluster-robust SE) | 116 (ESS: 47.5) | 28 (ESS: 10.2) | 2.52 ± 0.09 | 2.72 ± 0.13 | -2.057 | Cohen's d | <0.001 | <0.001 |
| Mean thickness of insula (left hemisphere) | Weighted t-test (Cluster-robust SE) | 116 (ESS: 47.5) | 28 (ESS: 10.2) | 2.75 ± 0.13 | 3.03 ± 0.21 | -1.917 | Cohen's d | <0.001 | <0.001 |
| Mean thickness of insula (right hemisphere) | Weighted t-test (Cluster-robust SE) | 116 (ESS: 47.5) | 28 (ESS: 10.2) | 2.70 ± 0.13 | 2.99 ± 0.16 | -2.177 | Cohen's d | <0.001 | <0.001 |
| Mean thickness of isthmuscingulate (left hemisphere) | Weighted t-test (Cluster-robust SE) | 116 (ESS: 47.5) | 28 (ESS: 10.2) | 2.07 ± 0.12 | 2.24 ± 0.14 | -1.395 | Cohen's d | <0.001 | <0.001 |
| Mean thickness of isthmuscingulate (right hemisphere) | Weighted t-test (Cluster-robust SE) | 116 (ESS: 47.5) | 28 (ESS: 10.2) | 2.06 ± 0.11 | 2.22 ± 0.11 | -1.454 | Cohen's d | <0.001 | <0.001 |
| Mean thickness of lateraloccipital (left hemisphere) | Weighted t-test (Cluster-robust SE) | 116 (ESS: 47.5) | 28 (ESS: 10.2) | 2.09 ± 0.09 | 2.20 ± 0.10 | -1.219 | Cohen's d | <0.001 | <0.001 |
| Mean thickness of lateraloccipital (right hemisphere) | Weighted t-test (Cluster-robust SE) | 116 (ESS: 47.5) | 28 (ESS: 10.2) | 2.15 ± 0.09 | 2.25 ± 0.14 | -1.054 | Cohen's d | 0.033 | 0.036 |
| Mean thickness of lateralorbitofrontal (left hemisphere) | Weighted t-test (Cluster-robust SE) | 116 (ESS: 47.5) | 28 (ESS: 10.2) | 2.39 ± 0.09 | 2.54 ± 0.11 | -1.571 | Cohen's d | <0.001 | <0.001 |
| Mean thickness of lateralorbitofrontal (right hemisphere) | Weighted t-test (Cluster-robust SE) | 116 (ESS: 47.5) | 28 (ESS: 10.2) | 2.31 ± 0.08 | 2.43 ± 0.11 | -1.352 | Cohen's d | 0.001 | 0.002 |
| Mean thickness of lingual (left hemisphere) | Weighted t-test (Cluster-robust SE) | 116 (ESS: 47.5) | 28 (ESS: 10.2) | 1.98 ± 0.08 | 2.14 ± 0.11 | -1.863 | Cohen's d | <0.001 | <0.001 |
| Mean thickness of lingual (right hemisphere) | Weighted t-test (Cluster-robust SE) | 116 (ESS: 47.5) | 28 (ESS: 10.2) | 1.98 ± 0.09 | 2.09 ± 0.09 | -1.212 | Cohen's d | <0.001 | 0.001 |
| Mean thickness of medialorbitofrontal (left hemisphere) | Weighted t-test (Cluster-robust SE) | 116 (ESS: 47.5) | 28 (ESS: 10.2) | 2.24 ± 0.10 | 2.33 ± 0.16 | -0.811 | Cohen's d | 0.086 | 0.09 |
| Mean thickness of medialorbitofrontal (right hemisphere) | Weighted t-test (Cluster-robust SE) | 116 (ESS: 47.5) | 28 (ESS: 10.2) | 2.32 ± 0.08 | 2.45 ± 0.13 | -1.414 | Cohen's d | 0.003 | 0.003 |
| Mean thickness of middletemporal (left hemisphere) | Weighted t-test (Cluster-robust SE) | 116 (ESS: 47.5) | 28 (ESS: 10.2) | 2.42 ± 0.09 | 2.62 ± 0.10 | -2.366 | Cohen's d | <0.001 | <0.001 |
| Mean thickness of middletemporal (right hemisphere) | Weighted t-test (Cluster-robust SE) | 116 (ESS: 47.5) | 28 (ESS: 10.2) | 2.43 ± 0.10 | 2.65 ± 0.09 | -2.276 | Cohen's d | <0.001 | <0.001 |
| Mean thickness of paracentral (left hemisphere) | Weighted t-test (Cluster-robust SE) | 116 (ESS: 47.5) | 28 (ESS: 10.2) | 2.34 ± 0.11 | 2.47 ± 0.16 | -1.077 | Cohen's d | 0.005 | 0.005 |
| Mean thickness of paracentral (right hemisphere) | Weighted t-test (Cluster-robust SE) | 116 (ESS: 47.5) | 28 (ESS: 10.2) | 2.27 ± 0.12 | 2.42 ± 0.21 | -1.15 | Cohen's d | 0.032 | 0.036 |
| Mean thickness of parahippocampal (left hemisphere) | Weighted t-test (Cluster-robust SE) | 116 (ESS: 47.5) | 28 (ESS: 10.2) | 2.45 ± 0.16 | 2.56 ± 0.30 | -0.571 | Cohen's d | 0.261 | 0.266 |
| Mean thickness of parahippocampal (right hemisphere) | Weighted t-test (Cluster-robust SE) | 116 (ESS: 47.5) | 28 (ESS: 10.2) | 2.31 ± 0.17 | 2.65 ± 0.16 | -2.05 | Cohen's d | <0.001 | <0.001 |
| Mean thickness of parsopercularis (left hemisphere) | Weighted t-test (Cluster-robust SE) | 116 (ESS: 47.5) | 28 (ESS: 10.2) | 2.29 ± 0.08 | 2.43 ± 0.14 | -1.397 | Cohen's d | 0.005 | 0.006 |
| Mean thickness of parsopercularis (right hemisphere) | Weighted t-test (Cluster-robust SE) | 116 (ESS: 47.5) | 28 (ESS: 10.2) | 2.25 ± 0.08 | 2.43 ± 0.08 | -2.297 | Cohen's d | <0.001 | <0.001 |
| Mean thickness of parsorbitalis (left hemisphere) | Weighted t-test (Cluster-robust SE) | 116 (ESS: 47.5) | 28 (ESS: 10.2) | 2.32 ± 0.11 | 2.52 ± 0.16 | -1.696 | Cohen's d | <0.001 | <0.001 |
| Mean thickness of parsorbitalis (right hemisphere) | Weighted t-test (Cluster-robust SE) | 116 (ESS: 47.5) | 28 (ESS: 10.2) | 2.32 ± 0.13 | 2.56 ± 0.15 | -1.86 | Cohen's d | <0.001 | <0.001 |
| Mean thickness of parstriangularis (left hemisphere) | Weighted t-test (Cluster-robust SE) | 116 (ESS: 47.5) | 28 (ESS: 10.2) | 2.14 ± 0.10 | 2.28 ± 0.10 | -1.431 | Cohen's d | <0.001 | <0.001 |
| Mean thickness of parstriangularis (right hemisphere) | Weighted t-test (Cluster-robust SE) | 116 (ESS: 47.5) | 28 (ESS: 10.2) | 2.13 ± 0.10 | 2.33 ± 0.09 | -2.077 | Cohen's d | <0.001 | <0.001 |
| Mean thickness of pericalcarine (left hemisphere) | Weighted t-test (Cluster-robust SE) | 116 (ESS: 47.5) | 28 (ESS: 10.2) | 1.63 ± 0.12 | 1.78 ± 0.13 | -1.193 | Cohen's d | <0.001 | 0.001 |
| Mean thickness of pericalcarine (right hemisphere) | Weighted t-test (Cluster-robust SE) | 116 (ESS: 47.5) | 28 (ESS: 10.2) | 1.62 ± 0.11 | 1.66 ± 0.11 | -0.347 | Cohen's d | 0.284 | 0.284 |
| Mean thickness of postcentral (left hemisphere) | Weighted t-test (Cluster-robust SE) | 116 (ESS: 47.5) | 28 (ESS: 10.2) | 1.98 ± 0.08 | 2.07 ± 0.09 | -1.055 | Cohen's d | 0.002 | 0.003 |
| Mean thickness of postcentral (right hemisphere) | Weighted t-test (Cluster-robust SE) | 116 (ESS: 47.5) | 28 (ESS: 10.2) | 1.96 ± 0.09 | 2.01 ± 0.09 | -0.512 | Cohen's d | 0.112 | 0.116 |
| Mean thickness of posteriorcingulate (left hemisphere) | Weighted t-test (Cluster-robust SE) | 116 (ESS: 47.5) | 28 (ESS: 10.2) | 2.10 ± 0.11 | 2.33 ± 0.14 | -1.911 | Cohen's d | <0.001 | <0.001 |
| Mean thickness of posteriorcingulate (right hemisphere) | Weighted t-test (Cluster-robust SE) | 116 (ESS: 47.5) | 28 (ESS: 10.2) | 2.11 ± 0.11 | 2.32 ± 0.11 | -1.975 | Cohen's d | <0.001 | <0.001 |
| Mean thickness of precentral (left hemisphere) | Weighted t-test (Cluster-robust SE) | 116 (ESS: 47.5) | 28 (ESS: 10.2) | 2.45 ± 0.10 | 2.58 ± 0.13 | -1.136 | Cohen's d | 0.001 | 0.001 |
| Mean thickness of precentral (right hemisphere) | Weighted t-test (Cluster-robust SE) | 116 (ESS: 47.5) | 28 (ESS: 10.2) | 2.40 ± 0.13 | 2.56 ± 0.12 | -1.236 | Cohen's d | <0.001 | <0.001 |
| Mean thickness of precuneus (left hemisphere) | Weighted t-test (Cluster-robust SE) | 116 (ESS: 47.5) | 28 (ESS: 10.2) | 2.19 ± 0.09 | 2.38 ± 0.08 | -2.268 | Cohen's d | <0.001 | <0.001 |
| Mean thickness of precuneus (right hemisphere) | Weighted t-test (Cluster-robust SE) | 116 (ESS: 47.5) | 28 (ESS: 10.2) | 2.18 ± 0.08 | 2.39 ± 0.08 | -2.55 | Cohen's d | <0.001 | <0.001 |
| Mean thickness of rostralanteriorcingulate (left hemisphere) | Weighted t-test (Cluster-robust SE) | 116 (ESS: 47.5) | 28 (ESS: 10.2) | 2.35 ± 0.16 | 2.52 ± 0.15 | -1.064 | Cohen's d | 0.003 | 0.003 |
| Mean thickness of rostralanteriorcingulate (right hemisphere) | Weighted t-test (Cluster-robust SE) | 116 (ESS: 47.5) | 28 (ESS: 10.2) | 2.39 ± 0.16 | 2.57 ± 0.15 | -1.137 | Cohen's d | <0.001 | <0.001 |
| Mean thickness of rostralmiddlefrontal (left hemisphere) | Weighted t-test (Cluster-robust SE) | 116 (ESS: 47.5) | 28 (ESS: 10.2) | 2.14 ± 0.08 | 2.26 ± 0.10 | -1.386 | Cohen's d | <0.001 | <0.001 |
| Mean thickness of rostralmiddlefrontal (right hemisphere) | Weighted t-test (Cluster-robust SE) | 116 (ESS: 47.5) | 28 (ESS: 10.2) | 2.11 ± 0.08 | 2.26 ± 0.08 | -1.883 | Cohen's d | <0.001 | <0.001 |
| Mean thickness of superiorfrontal (left hemisphere) | Weighted t-test (Cluster-robust SE) | 116 (ESS: 47.5) | 28 (ESS: 10.2) | 2.39 ± 0.09 | 2.51 ± 0.11 | -1.28 | Cohen's d | <0.001 | <0.001 |
| Mean thickness of superiorfrontal (right hemisphere) | Weighted t-test (Cluster-robust SE) | 116 (ESS: 47.5) | 28 (ESS: 10.2) | 2.36 ± 0.07 | 2.53 ± 0.08 | -2.498 | Cohen's d | <0.001 | <0.001 |
| Mean thickness of superiorparietal (left hemisphere) | Weighted t-test (Cluster-robust SE) | 116 (ESS: 47.5) | 28 (ESS: 10.2) | 2.09 ± 0.09 | 2.24 ± 0.08 | -1.632 | Cohen's d | <0.001 | <0.001 |
| Mean thickness of superiorparietal (right hemisphere) | Weighted t-test (Cluster-robust SE) | 116 (ESS: 47.5) | 28 (ESS: 10.2) | 2.06 ± 0.09 | 2.19 ± 0.09 | -1.399 | Cohen's d | <0.001 | <0.001 |
| Mean thickness of superiortemporal (left hemisphere) | Weighted t-test (Cluster-robust SE) | 116 (ESS: 47.5) | 28 (ESS: 10.2) | 2.50 ± 0.09 | 2.70 ± 0.13 | -2.054 | Cohen's d | <0.001 | <0.001 |
| Mean thickness of superiortemporal (right hemisphere) | Weighted t-test (Cluster-robust SE) | 116 (ESS: 47.5) | 28 (ESS: 10.2) | 2.49 ± 0.09 | 2.75 ± 0.14 | -2.512 | Cohen's d | <0.001 | <0.001 |
| Mean thickness of supramarginal (left hemisphere) | Weighted t-test (Cluster-robust SE) | 116 (ESS: 47.5) | 28 (ESS: 10.2) | 2.26 ± 0.08 | 2.47 ± 0.08 | -2.44 | Cohen's d | <0.001 | <0.001 |
| Mean thickness of supramarginal (right hemisphere) | Weighted t-test (Cluster-robust SE) | 116 (ESS: 47.5) | 28 (ESS: 10.2) | 2.25 ± 0.09 | 2.46 ± 0.09 | -2.441 | Cohen's d | <0.001 | <0.001 |
| Mean thickness of transversetemporal (left hemisphere) | Weighted t-test (Cluster-robust SE) | 116 (ESS: 47.5) | 28 (ESS: 10.2) | 2.19 ± 0.15 | 2.37 ± 0.20 | -1.189 | Cohen's d | 0.004 | 0.005 |
| Mean thickness of transversetemporal (right hemisphere) | Weighted t-test (Cluster-robust SE) | 116 (ESS: 47.5) | 28 (ESS: 10.2) | 2.21 ± 0.14 | 2.42 ± 0.20 | -1.39 | Cohen's d | 0.004 | 0.005 |
| Note: All tests are Weighted t-tests with Cluster-robust standard errors. Effective sample sizes are reported in parenthesis. Abbreviations: ESS, Effective Sample Size | | | | | | | | | |

**Table S10. Comparisons on outcome variables between clusters from the HSR validation cohort**

| **Variable** | **Cluster 1 (N=116)** | **Cluster (N=28)** | **ES** | **p** | **q** |
| --- | --- | --- | --- | --- | --- |
| Scanner (% Ingenia CX) | 74/116 (63.8%) | 18/28 (64.3%) | V = 0.000 | 1 | 1 |
| Age (years) | 52.14 ± 8.80 | 46.32 ± 11.92 | d = 0.614 | 0.021 | 0.242 |
| Sex (% male) | 46/116 (39.7%) | 9/28 (32.1%) | V = 0.043 | 0.605 | 0.851 |
| BMI | 25.39 ± 4.94 | 24.19 ± 4.69 | d = 0.244 | 0.256 | 0.815 |
| Education | 13.00 (8.00–17.00) | 13.00 (8.00–16.50) | d = -0.073 | 0.776 | 0.892 |
| Age of onset (years) | 33.83 ± 11.87 | 28.86 ± 8.67 | d = 0.439 | 0.015 | 0.242 |
| Number of depressive episodes | 4.00 (2.00–6.25) | 3.00 (2.00–5.25) | d = 0.219 | 0.48 | 0.815 |
| Pharmacological load | 4.00 (3.00–6.00) | 4.00 (3.00–7.00) | d = -0.136 | 0.722 | 0.892 |
| Cardiometabolic comorbidity | 13/116 (11.2%) | 0/28 (0.0%) | V = 0.124 | 0.073 | 0.426 |
| Depression with atypical features | 2/112 (1.8%) | 1/28 (3.6%) | V = 0.000 | 0.491 | 0.815 |
| Depression with anxious features | 22/112 (19.6%) | 5/28 (17.9%) | V = 0.000 | 1 | 1 |
| Treatment resistant depression | 47/112 (42.0%) | 6/28 (21.4%) | V = 0.151 | 0.074 | 0.426 |
| HDRS-21 total score | 22.75 ± 7.00 | 21.93 ± 6.71 | d = 0.119 | 0.575 | 0.851 |
| BDI total score | 16.38 ± 7.79 | 17.95 ± 6.63 | d = -0.207 | 0.421 | 0.815 |
| BDI Negative Self-Esteem | 5.38 ± 3.35 | 5.95 ± 2.66 | d = -0.175 | 0.496 | 0.815 |
| BDI Anergy | 5.68 ± 2.59 | 5.89 ± 2.81 | d = -0.080 | 0.756 | 0.892 |
| BDI Dysphoria | 5.47 ± 2.96 | 6.16 ± 2.57 | d = -0.237 | 0.358 | 0.815 |
| CTQ total score | 41.12 ± 11.82 | 44.27 ± 11.95 | d = -0.266 | 0.267 | 0.815 |
| CTQ Physical Abuse | 5.00 (5.00–6.00) | 5.00 (5.00–6.00) | d = 0.033 | 0.629 | 0.851 |
| CTQ Emotional Neglect | 13.10 ± 5.03 | 14.27 ± 5.59 | d = -0.227 | 0.344 | 0.815 |
| CTQ Emotional Abuse | 7.00 (5.00–10.00) | 8.00 (5.00–10.75) | d = -0.227 | 0.479 | 0.815 |
| CTQ Physical Neglect | 6.00 (5.00–9.00) | 6.00 (5.00–8.75) | d = -0.090 | 0.909 | 0.996 |
| CTQ Sexual Abuse | 5.00 (5.00–5.00) | 5.00 (5.00–5.75) | d = -0.219 | 0.143 | 0.658 |
| Abbreviations: ES, Effect Size; BMI, Body Mass Index; HDRS-21, Hamilton Depression Rating Scale; BDI, Beck Depression Inventory; CTQ, Childhood Trauma Questionnaire. | | | | | |

**Table S11. Summary of subsets of matched healhty controls derived from greedy matching algorithm**

| **subset id** | **n controls** | **worst pvalue** | **p Age** | **p Sex** |
| --- | --- | --- | --- | --- |
| 1 | 827 | 0.0361 | 0.0361 | 0.6937 |
| 2 | 816 | 0.0375 | 0.0375 | 0.6777 |
| 3 | 794 | 0.0361 | 0.0361 | 0.6784 |
| 4 | 791 | 0.0361 | 0.0361 | 0.6798 |
| 5 | 778 | 0.0347 | 0.0347 | 0.6784 |
| 6 | 778 | 0.0347 | 0.0347 | 0.6784 |
| 7 | 777 | 0.0368 | 0.0368 | 0.6895 |
| 8 | 771 | 0.0368 | 0.0368 | 0.6861 |
| 9 | 768 | 0.0382 | 0.0382 | 0.6840 |
| 10 | 768 | 0.0375 | 0.0375 | 0.6840 |
| 11 | 742 | 0.0368 | 0.0368 | 0.6923 |
| 12 | 711 | 0.0354 | 0.0354 | 0.6861 |
| 13 | 633 | 0.0389 | 0.03889 | 0.5576 |
| 14 | 598 | 0.0375 | 0.0375 | 0.6299 |
| 15 | 593 | 0.0375 | 0.0375 | 0.6881 |

**Table S12. List of outcome variables included in UKB and HSR cohorts**

|  | **UKB** |  |  | **HSR** |  |  |
| --- | --- | --- | --- | --- | --- | --- |
| Variable name | UKB Variable name (UKB Data-Field) | Coding | Type | HSR Variable name | Coding | Type |
| Socio-demographic variables |  |  |  |  |  |  |
| Age | Age when attended assessment centre \| Instance 02 (21003) | Age in years | Numeric | Age at the time of recruitment | Age in years | Numeric |
| Sex | Sex (31) | 1=Male; 0=Female | Binary | Sex | 1=Male; 0=Female | Binary |
| Education | Qualifications \| Instance 02 (6138) | Each number correpsond to a distinct categories of educational level. 1=CSEs; 2=O levels / GCSEs; 3=NVQ or HND or HNC; 4=A levels / AS levels; 5=Other professional qualifications; 6=College or University degree; NA=None of the above / Prefer not to answer | Categorical | Years of education | Education in years | Numeric |
| Depression-related variables |  |  |  |  |  |  |
| Depressive symptomatology | Recent feelings of depression \| Instance 02 (20510); Recent feelings of inadequacy \| Instance 02 (20507); Recent feelings of tiredness or low energy \| Instance 02 (20519); Recent lack of interest or pleasure in doing things \| Instance 02 (20514) | 0=Not at all; 1=Several days; 2=More than half the days; 3=Nearly every day; NA=None of the above / Prefer not to answer | Categorical | Total score of Hamilton Depression Rating Scale - 21 Items; Total score of Beck Depression Rating Scale - Short Form; BDI-SF Domains of Anergy, Negative Self-Esteem, and Dysphoria derived from doi: 10.1002/1097-4679(198701)43:1<111::aid-jclp2270430118>3.0.co;2-s. | Total scores and domain scores | Numeric |
| Age of onset | Difference in years between the date of first reported depressive episode (130894; 130896) and age when attended assessment centre \| Instance 02 (21003) | Age in years | Numeric | Age of onset of first depressive episode | Age in years | Numeric |
| Number of depressive episodes | Number of depression episodes \| Instance 02 (4620) | Number of episodes | Numeric | Number of depressive episodes | Number of episodes | Numeric |
| Depression with anxious features | Deived from https://doi.org/10.1038%2Fs41386-021-01059-6 | 1=Yes; 0=No | Binary | Depression with anxious features (DSM-5 specifier) | 1=Yes; 0=No | Binary |
| Depression with atypical features | Derived from https://doi.org/10.1038%2Fs41386-021-01059-6 | 1=Yes; 0=No | Binary | Depression with atypical features (DSM-5 specifier) | 1=Yes; 0=No | Binary |
| Antidepressant treatment | Dichotomous antidepressant treatment indicator | 1=Yes; 0=No | Binary | Pharnacological load defined according to Sackeim (2001) (PMID: 11480879) | Individual pharmacological load | Numeric |
| Treatment resistant depression | Derived from 10.1038/s41380-021-01062-9 | 1=Yes; 0=No | Binary | Derived from Thase & Rush criteria (PMID: 9402916) | 1=Yes; 0=No | Binary |
| Childhood trauma |  |  |  |  |  |  |
| Emotional Neglect | Felt love as a child (20489) | 0=Never; 1=Rarely true; 2=Sometimes true; 3=Often; 4=very often | Categorical | CTQ Emotional Neglect domain | Derived from https://doi.org/10.1016/S0145-2134(02)00541-0 | Numeric |
| Emotional Abuse | Felt hated by family member as a child (20487) | 0=Never; 1=Rarely true; 2=Sometimes true; 3=Often; 4=very often | Categorical | CTQ Emotional Abuse domain | Derived from https://doi.org/10.1016/S0145-2134(02)00541-0 | Numeric |
| Physical Abuse | Physically abused by family as a child (20488) | 0=Never; 1=Rarely true; 2=Sometimes true; 3=Often; 4=very often | Categorical | CTQ Physical Abuse domain | Derived from https://doi.org/10.1016/S0145-2134(02)00541-0 | Numeric |
| Sexual Abuse | Sexually molested as a child (20490) | 0=Never; 1=Rarely true; 2=Sometimes true; 3=Often; 4=very often | Categorical | CTQ Sexual Abuse domain | Derived from https://doi.org/10.1016/S0145-2134(02)00541-0 | Numeric |
| Physical Neglect | Someone to take to doctor when needed as a child (20491) | 0=Never; 1=Rarely true; 2=Sometimes true; 3=Often; 4=very often | Categorical | CTQ Physical Neglect domain | Derived from https://doi.org/10.1016/S0145-2134(02)00541-0 | Numeric |
| Childhood trauma exposure | Sum of 20489 (reverse), 20487, 20488, 20490, and 20491 (reverse) | Total score | Numeric | CTQ total score | Sum of CTQ domains | Numeric |
| Health-related variables |  |  |  |  |  |  |
| Body mass index (BMI) | Body mass index \| Instance 02 (23104) | Value in Kg/m2 | Numeric | Body mass index at the recruitment | Value in Kg/m2 | Numeric |
| Diabetes / cardiometabolic comorbidity | Diabetes diagnosed by a doctor \| Instance 02 (2443) | 1=Yes; 0=No; NA=Do not know / Prefer not to answer | Binary | Presence of cardiometabolic comorbidity in anamnesis | 1=Yes; 0=No | Binary |

**Table S13. List of drugs included in antidepressant medications in HSR cohort**

**List of antidepressant medications**

Venlafaxine

Escitalopram

Citalopram

Sertraline

Mirtazapine

Trazodone

Fluvoxamine

Fluoxetine

Duloxetine

Paroxetine

Clomipramine

Imipramine

Nortriptyline

Trimipramine

Vortioxetine

Bupropion

Amitriptyline

**Table S14. Neurosynth terms**

| **action** | **face recognition** | **motor control** | **strategy** |
| --- | --- | --- | --- |
| adaptation | facial expression | movement | strength |
| addiction | familiarity | multisensory | stress |
| anticipation | fear | naming | sustained attention |
| anxiety | fixation | navigation | task difficulty |
| arousal | focus | object recognition | thought |
| association | gaze | pain | uncertainty |
| attention | goal | perception | updating |
| autobiographical memory | hyperactivity | planning | utility |
| balance | imagery | priming | valence |
| belief | impulsivity | psychosis | verbal fluency |
| categorization | induction | reading | visual attention |
| cognitive control | inference | reasoning | visual perception |
| communication | inhibition | recall | word recognition |
| competition | insight | recognition | working memory |
| concept | integration | rehearsal |  |
| consciousness | intelligence | reinforcement learning |  |
| consolidation | intention | response inhibition |  |
| context | interference | response selection |  |
| coordination | judgment | retention |  |
| decision | knowledge | retrieval |  |
| decision making | language | reward anticipation |  |
| detection | language comprehension | rhythm |  |
| discrimination | learning | risk |  |
| distraction | listening | rule |  |
| eating | localization | salience |  |
| efficiency | loss | search |  |
| effort | maintenance | selective attention |  |
| emotion | manipulation | semantic memory |  |
| emotion regulation | meaning | sentence comprehension |  |
| empathy | memory | skill |  |
| encoding | memory retrieval | sleep |  |
| episodic memory | mental imagery | social cognition |  |
| expectancy | monitoring | spatial attention |  |
| expertise | mood | speech perception |  |
| extinction | morphology | speech production |  |

**Table S15. Neurotransmitter receptors and transporters used to build the receptor density matrix.**

| **Receptor/transporter** | **Neurotransmitter** | **Tracer** | **Measure** | **#Participants** | **Age (years)** | **References** |
| --- | --- | --- | --- | --- | --- | --- |
| 5-HT₁ₐ | serotonin | [¹¹C]WAY-100635 | BPND | 35 (17) | 26.3 ± 5.2 | Savli et al., 2012 |
| 5-HT₁ᵦ | serotonin | [¹¹C]P943 | BPND | 23 (8) | 28.7 ± 7.0 | Savli et al., 2012 |
| 5-HT₁ᵦ | serotonin | [¹¹C]P943 | BPND | 65 (16) | 33.7 ± 9.7 | Gallezot et al., 2010 |
| 5-HT₂ₐ | serotonin | [¹¹C]Cimbi-36 | Bmax | 29 (14) | 22.6 ± 2.7 | Beliveau et al., 2017 |
| 5-HT₄ | serotonin | [¹¹C]SB207145 | Bmax | 59 (18) | 25.9 ± 5.3 | Beliveau et al., 2017 |
| 5-HT₆ | serotonin | [¹¹C]GSK215083 | BPND | 30 (0) | 36.6 ± 9.0 | Radhakrishnan et al., 2018 |
| 5-HTT* | serotonin | [¹¹C]DASB | Bmax | 100 (71) | 25.1 ± 5.8 | Beliveau et al., 2017 |
| α4β2 | acetylcholine | [¹⁸F]FLUBATINE | VT | 30 (10) | 33.5 ± 10.7 | Hillmer et al., 2016 |
| CB₁ | cannabinoid | [¹¹C]OMAR | VT | 77 (28) | 30.0 ± 8.9 | Normandin et al., 2015 |
| D₁ | dopamine | [¹¹C]SCH23390 | BPND | 13 (7) | 33 ± 13 | Kaller et al., 2017 |
| D₂ | dopamine | [¹¹C]FLB-457 | BPND | 37 (20) | 48.4 ± 16.9 | Smith et al., 2019 |
| D₂ | dopamine | [¹¹C]FLB-457 | BPND | 55 (29) | 32.5 ± 9.7 | Sandiego et al., 2015 |
| DAT* | dopamine | [¹²³I]-FP-CIT | SUVR | 174 (65) | 61 ± 11 | Dukart et al., 2018 |
| GABAA/BZ | GABA | [¹¹C]flumazenil | Bmax | 16 (9) | 26.6 ± 8 | Nørgaard et al., 2021 |
| H₃ | histamine | [¹¹C]GSK189254 | VT | 8 (1) | 31.7 ± 9.0 | Gallezot et al., 2017 |
| M₁ | acetylcholine | [¹¹C]LSN3172176 | BPND | 24 (11) | 40.5 ± 11.7 | Naganawa et al., 2021 |
| mGluR₅ | glutamate | [¹¹C]ABP688 | BPND | 22 (10) | 67.9 ± 9.6 | Rosa-Neto & Kobayashi |
| mGluR₅ | glutamate | [¹¹C]ABP688 | BPND | 28 (13) | 33.1 ± 11.2 | Dubois et al., 2016 |
| mGluR₅ | glutamate | [¹¹C]ABP688 | BPND | 73 (48) | 19.9 ± 3.0 | Smart et al., 2019 |
| MOR | opioid | [¹¹C]carfentanil | BPND | 204 (72) | 32.3 ± 10.8 | Kantonen et al., 2020 |
| NET* | norepinephrine | [¹¹C]MRB | BPND | 77 (27) | 33.4 ± 9.2 | Ding et al., 2010 |
| NMDAR | glutamate | [¹⁸F]GE-179 | VT | 29 (8) | 40.9 ± 12.7 | Galovic et al., 2021 |
| VAChT* | acetylcholine | [¹⁸F]FEOBV | SUVR | 4 (1) | 37 ± 10.2 | Tuominen & Guimond |
| VAChT* | acetylcholine | [¹⁸F]FEOBV | SUVR | 5 (1) | 68.3 ± 3.1 | Bédard et al., 2019 |
| VAChT* | acetylcholine | [¹⁸F]FEOBV | SUVR | 18 (13) | 66.8 ± 6.8 | Aghourian et al., 2017 |
| Abbreviations, BPND, non-displaceable binding potential; VT, tracer distribution volume; Bmax, density (pmol/ml) converted from binding potential (5-HT) or distributional volume (GABA) using autoradiography-derived densities; SUVR, standard uptake value ratio. Values in parentheses under #Participants indicate number of females. Neurotransmitter receptor maps without citations refer to previously unpublished data. In those cases, contact information for the study principal investigator is provided in the Table. Asterisks indicate transporters. | | | | | | |
